# What is the current state of public health system preparedness for infectious disease emergencies? A scoping review

**DOI:** 10.1101/2022.10.25.22281308

**Authors:** Jessica M Lee, Rachel Jansen, Kate E Sanderson, Fiona Guerra, Sue Keller-Olaman, Michelle Murti, Tracey L O’Sullivan, Madelyn P Law, Brian Schwartz, Laura E Bourns, Yasmin Khan

## Abstract

**Background:** The COVID-19 pandemic continues to demonstrate the risks and profound health impacts that result from infectious disease emergencies. Emergency preparedness has been defined as the knowledge, capacity and organizational systems that governments, response and recovery organizations, communities and individuals develop to anticipate, respond to, or recover from emergencies. This scoping review explored recent literature on priority areas and indicators for public health emergency preparedness (PHEP) with a focus on infectious disease emergencies.

**Methods:** Using scoping review methodology, a comprehensive search was conducted for indexed and grey literature with a focus on records published from 2017 and 2020 onward, respectively. Records were included if they: a) described PHEP, b) focused on an infectious emergency, and c) were published in an Organization for Economic Co-operation and Development country. An evidence-based all-hazards Resilience Framework for PHEP consisting of 11 elements was used as a reference point to identify additional areas of preparedness that have emerged in recent publications. The findings were summarized thematically.

**Results:** The included publications largely aligned with the all-hazards Resilience Framework for PHEP. In particular, the elements related to collaborative networks, community engagement, risk analysis and communication were frequently observed across the publications included in this review. Emergent themes were identified that expand on the Resilience Framework for PHEP. These were related to mitigating inequities, public health capacities (vaccination, laboratory system capacity, infection prevention and control capacity, financial investment in infrastructure, public health legislation, phases of preparedness), scientific capacities (research and evidence-informed decision making, climate and environmental health), and considerations for health system capacity.

**Conclusions:** The themes from this review contribute to the evolving understanding of critical public health preparedness actions; however, there was a paucity of recent evidence on PHEP indicators. The themes can expand on the 11 elements outlined in the Resilience Framework for PHEP, specifically relevant to infectious disease emergencies and risks. Further research will be important to validate these findings, and expand understanding of how refinements to PHEP frameworks and indicators can support public health practice.

## 1.0 Background

The Coronavirus Disease 2019 (COVID-19) pandemic is responsible for millions of deaths globally [1, 2], and continues to demonstrate the risks and profound health impacts that result from infectious disease emergencies. While the impacts of disasters and emergencies were known to have inequitable impacts across populations prior to the COVID-19 pandemic [3, 4], the disparities in COVID-19 outcomes have been grave [5-7]. Since the start of the pandemic, there have been demonstrable inequities in COVID-19 morbidity and mortality in marginalized communities such as racialized, low-income and Indigenous communities in Canada, as well as inequitable impacts of implementing and removing public health measures at different time periods throughout the pandemic [7-9]. Ecological impacts of climate change, population growth trends, and increasing population density are amongst the factors increasing global risks for the emergence of novel infectious diseases [10-12]. It is crucial to ensure a continued review and reflection on emergency preparedness to assess ongoing risks, to reduce morbidity and mortality, and to mitigate the inequitable impacts of infectious disease emergencies and response measures, which is the focus of this paper.

Following the 2003 Severe Acute Respiratory Syndrome (i.e., SARS-CoV-1) outbreak and the H1N1 influenza pandemic, there was a lack of evidence to inform defining and measuring public health emergency preparedness (PHEP) [13]. The World Health Organization (WHO) Strategic Framework (2017) has defined emergency preparedness as the knowledge, capacity and organizational system that governments, response and recovery organizations, communities, and individuals develop to anticipate, respond to, or recover from emergencies [14]. Operationally, emergency preparedness involves specific actions, funding, partnerships and political commitment to be sustainable [14]. Investing in and implementing priority actions requires an understanding of these characteristics and elements of preparedness. Sustaining preparedness actions can benefit from metrics to describe, assess, and report on change over time.

To address the knowledge gap in defining and measuring PHEP, a Canadian-based research team, including several authors of this paper, explored PHEP for infectious and non-infectious emergencies, and developed an evidence-based, all-hazards Resilience Framework for PHEP [15-17]. The framework consists of the following elements: governance and leadership (cross-cutting), planning process, collaborative networks, community engagement, risk analysis, surveillance and monitoring, practice and experience, resources, workforce capacity, communication, and learning and evaluation. The set of 67 important and actionable PHEP indicators correspond with the framework’s elements and can be used by public health agencies to assess readiness and measure improvement in their critical role of protecting community health. Ethics and values are included in this framework as a concept that should be considered as core to all elements of PHEP rather than as a specific element with corresponding indicators. While the evidence-based Resilience Framework for PHEP preceded the COVID-19 pandemic, it represented a novel contribution to the field which had limited evidence to inform practice on a framework and indicators for preparedness [15-17]. Given the global experience with the COVID-19 pandemic, there is value in exploring how the evidence base has developed since the framework and indicators were created, with a focus on infectious disease preparedness.

In this scoping review, we explored the literature on frameworks, priority areas and indicators for PHEP with a focus on infectious disease emergencies. We used the Resilience Framework for PHEP to examine areas of preparedness actions and indicators developed in the period since the previous study was conducted [15-17], which includes the COVID-19 pandemic period. The objective of this scoping review is to investigate the following two research questions:

1. What recent evidence has emerged on conceptual frameworks for PHEP specific to pandemics and infectious disease emergencies?
2. What recent evidence has emerged pertaining to measurement of preparedness for pandemics and infectious disease emergencies?

## 2.0 Methods

### 2.1 Aim and design

A scoping review methodology was used, given the exploratory nature of the research questions. This method focuses on mapping concepts underpinning a research area and is useful when examining areas that are emerging, to clarify key concepts and identify gaps [18-20]. Consistent with the scoping review methodology, a quality appraisal of the included studies was not conducted [18-20]. With this scoping review, we aimed to expand the understanding of the current state of PHEP frameworks, priority areas and indicators relevant to public health agencies, and how these may have evolved during the COVID-19 pandemic. The focus of this review was on local and/or regional or provincial/state (i.e. sub-national) public health, given that the public health system in Canada is organized around local/regional public health agencies, with provincial health system governance and organization [16, 17].

### 2.2 Data sources and search methods

Library information specialists at Public Health Ontario (PHO) were consulted to conduct database searches in MEDLINE (March 22, 2022); Embase, Business Continuity & Disaster Recovery Reference Center and Scopus (March 28, 2022); and the National Institutes of Health COVID-19 Portfolio for Preprints (March 15, 2022). The search included terms related to public health emergencies, emergency preparedness, post-pandemic recovery, indicators/measures, and frameworks.

The indexed literature search focused on identifying publications that included a description of frameworks, tools, models, activities or indicators for emergency preparedness for infectious diseases, pandemic influenza and the COVID-19 pandemic from 2017 onward. This approach captured literature published since the scoping review was conducted to inform the Delphi expert panel for PHEP indicator development [17], as well as literature published during the COVID-19 pandemic.

In addition to the indexed literature, a grey literature search was conducted from March 17, 2022 to March 25, 2022. PHO library information specialists were consulted to develop search strings to be used in Google Custom Search Engines and select regional, national and international public health agency websites. The search was limited to records published from 2020 onward to capture frameworks, models, toolkits and indicators published within the COVID-19 pandemic context. See additional file 1 for the full indexed and grey literature search strategies.

### 2.3 Eligibility criteria and record selection

The eligibility criteria were the same across indexed and grey literature except for the time periods searched, as noted above. Records were included in the scoping review if they met the following criteria: a) planning, readiness and preparedness included the roles and responsibilities of local, regional or provincial/state/sub-national public health agencies relevant to Canada; b) emergency described in the article or framework was a pandemic and/or of infectious origins; c) the emergency or framework described was specific to an Organization for Economic Co-operation and Development (OECD) country; and d) described preparedness activities, including indicators to inform preparedness activities, that are under local, regional or provincial/state (i.e. sub-national) level jurisdiction.

Records describing federal-, national-, or international-level (e.g., WHO) relevant frameworks or indicators were also included if the roles, responsibilities, elements and/or indicators described were relevant to public health agencies and public health system organization in Canada. For example, frameworks which described surveillance and laboratory testing activities were included, whereas a focus on measures relevant only to the federal level in Canada such as travel quarantine would be excluded. Infectious disease emergency was defined as an incident, outbreak or threat with the potential to overwhelm or otherwise disrupt routine local capacities due to their timing, scale or unpredictability [21-23]. Only English-language records were included.

Records were excluded if they: a) focused on non-preparedness components of emergency management (i.e., response, recovery and mitigation); b) described an emergency of non-infectious origins; c) described a framework limited to country or federal-level roles relevant to Canada and countries with similar health system organization (i.e., travel or international border measures); or d) focused on health care system (e.g., primary care, acute care) preparedness without public health system considerations [16]. Commentaries were excluded.

Results of the indexed literature search were screened by two authors. A random selection of 100 records were first screened independently in duplicate to check agreement and trial the eligibility criteria, which achieved 84% agreement. This allowed the two authors to discuss discrepancies and reach consensus on the articles, leading to enhanced understanding and consistency in how the remaining records were screened. Single author screening occurred for the remainder of indexed literature results, and a third author was consulted for uncertainties related to inclusion of specific studies when required. The grey literature search and screening were conducted by two authors. Similar to the process for indexed literature, a third author was consulted for uncertainties related to the inclusion of grey literature records when required.

### 2.4 Data extraction, summary and synthesis

The following details were extracted from the included publications: year, country/jurisdiction, relevant jurisdictional level (e.g., national, provincial, regional, or local/municipal), type of infectious disease emergency (i.e., COVID-19, any infectious disease), study and/or framework design and objective(s), description of the framework’s elements or components, and description of the framework indicators (if applicable). For records related to all-hazards PHEP, only details relevant to infectious diseases were extracted.

The first step in analysis was identifying emergent themes from the literature. For the purposes of synthesizing the results in this scoping review, we refer to the high-level topic areas related to public health preparedness for infectious disease emergencies as themes, which could be associated with related actions or indicators. Sources included in this report used various terms including “principles”, “domains”, “elements”, “dimensions”, “key areas”, “categories” and others. In this first step, the previous research by Khan et al. was used as a reference point for synthesizing PHEP elements and emergent themes we identified in the literature [16, 17]. Although Khan et al.’s work examined preparedness for all-hazards emergencies, the scope of this review was focused on identifying emerging themes related to public health preparedness for infectious disease emergencies, including pandemics. The elements of the Resilience Framework for PHEP were also used as a reference point to identify new areas of preparedness that have emerged in the literature. Ethics and values were considered as part of the 11 elements, rather than separate, consistent with the Resilience Framework for PHEP [15].

In the second step, identified themes were compared and contrasted with the elements of the Resilience Framework for PHEP to examine similarities and/or differences [15]. We identified elements of the Resilience Framework for PHEP that were described in the literature, and preparedness themes that have emerged since the previous scoping review and indicator development work of Khan, et al. [16, 17], and since the COVID-19 pandemic. Where alignment was observed, we report how frequently each of the 11 elements were observed across included publications. When themes did not overlap with the Resilience Framework for PHEP, these were recorded as “emergent themes” that expand upon the framework. The emergent themes were assessed for similarities, then synthesized into overarching topics. The indexed and grey literature were synthesized separately. Team members compared their lists of emergent themes, and where appropriate, aligned the language to describe common emergent themes. We also included examples of preparedness activities relevant to various PHEP elements and emergent themes.

The final step was to examine included studies for indicators or actions/activities that could be used to inform the development of indicators. Indicators were defined as succinct measures that help understand, compare and improve systems [24]; they are generally found in frameworks, assessment tools or checklists. For the purposes of this review, broad areas of measurement (e.g., vaccination) were synthesized rather than specific indicators (e.g., vaccinate specific proportion of the population) from a given framework or publication.

## 3.0 Results

From the 3,603 records identified through the peer-reviewed and pre-print literature database searches, 315 full-text records were assessed for eligibility and 26 studies were included in this scoping review. Of the records identified from searching organizational or government databases in the grey literature search, 179 were assessed for eligibility and 10 grey literature publications were included in this scoping review (see PRISMA diagram in figure 1). In summary, 36 records were examined for this scoping review.

**Figure 1.**
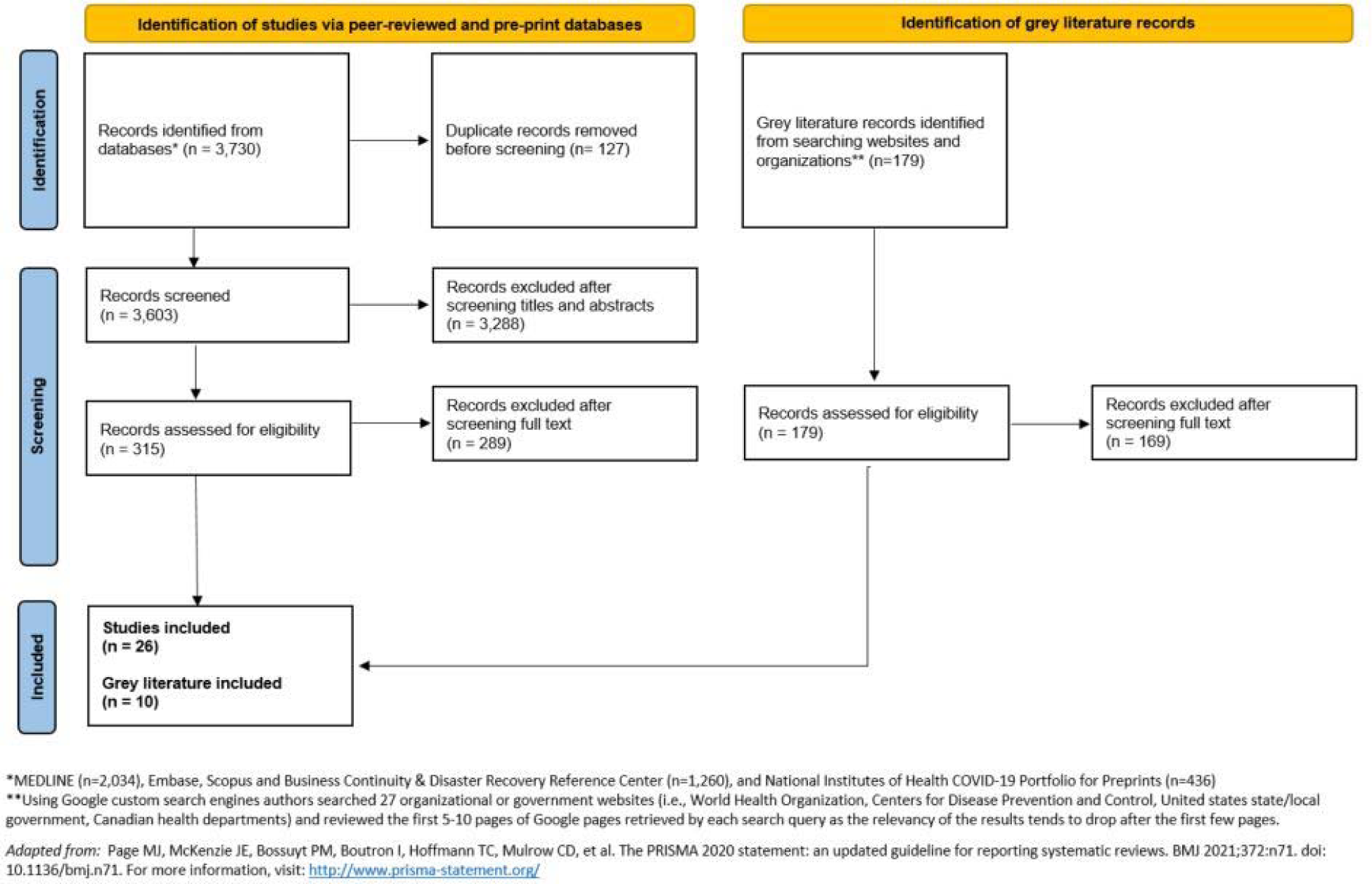
Flow chart of included records from indexed databases and grey literature searches.

### 3.1 Characteristics of included publications

Methods and study designs varied widely across the 26 indexed literature studies, including systematic literature reviews [25-27], mixed-methods studies (i.e., a paper that describes a literature review, concept mapping and key informant interviews) [28-43], descriptive case studies [44-46], qualitative studies [47, 48], a cross-sectional study [49], and a regression analysis [50]. Ten studies described a PHEP-related framework, tool or model [30-34, 36-39, 41], and 16 studies included content relevant to PHEP priority areas and/or activities but did not explicitly describe a PHEP framework, tool, model or set of indicators [25-29, 35, 40, 42-50]. All studies from the indexed literature described PHEP concepts for infectious disease outbreaks, pandemic influenza and/or the COVID-19 pandemic.

A total of 10 grey literature publications were identified, including four that described PHEP frameworks or conceptual models [51-54], three that described assessment tools [55-57], and three that focused on indicators for PHEP [58-60]. All ten grey literature publications described public health preparedness actions for infectious disease outbreaks, COVID-19 pandemic or zoonotic disease outbreaks. Of the ten documents identified, seven were produced by the WHO [51-57].

### 3.2 Elements from the Resilience Framework for PHEP that appeared in the included publications

After the first and second steps in analysis of the included studies, at least one element from the Resilience Framework for PHEP was observed in the 26 indexed studies [25-50], and seven of the ten grey literature records [51-57], with many studies making reference to multiple elements (see Table 1). The 11 elements, listed from most to least frequently observed across the included publications, were: collaborative networks, community engagement, risk analysis, communication, planning process, governance and leadership, surveillance and monitoring, resources, workforce capacity, learning and evaluation, and practice and experience.

**Table 1.**
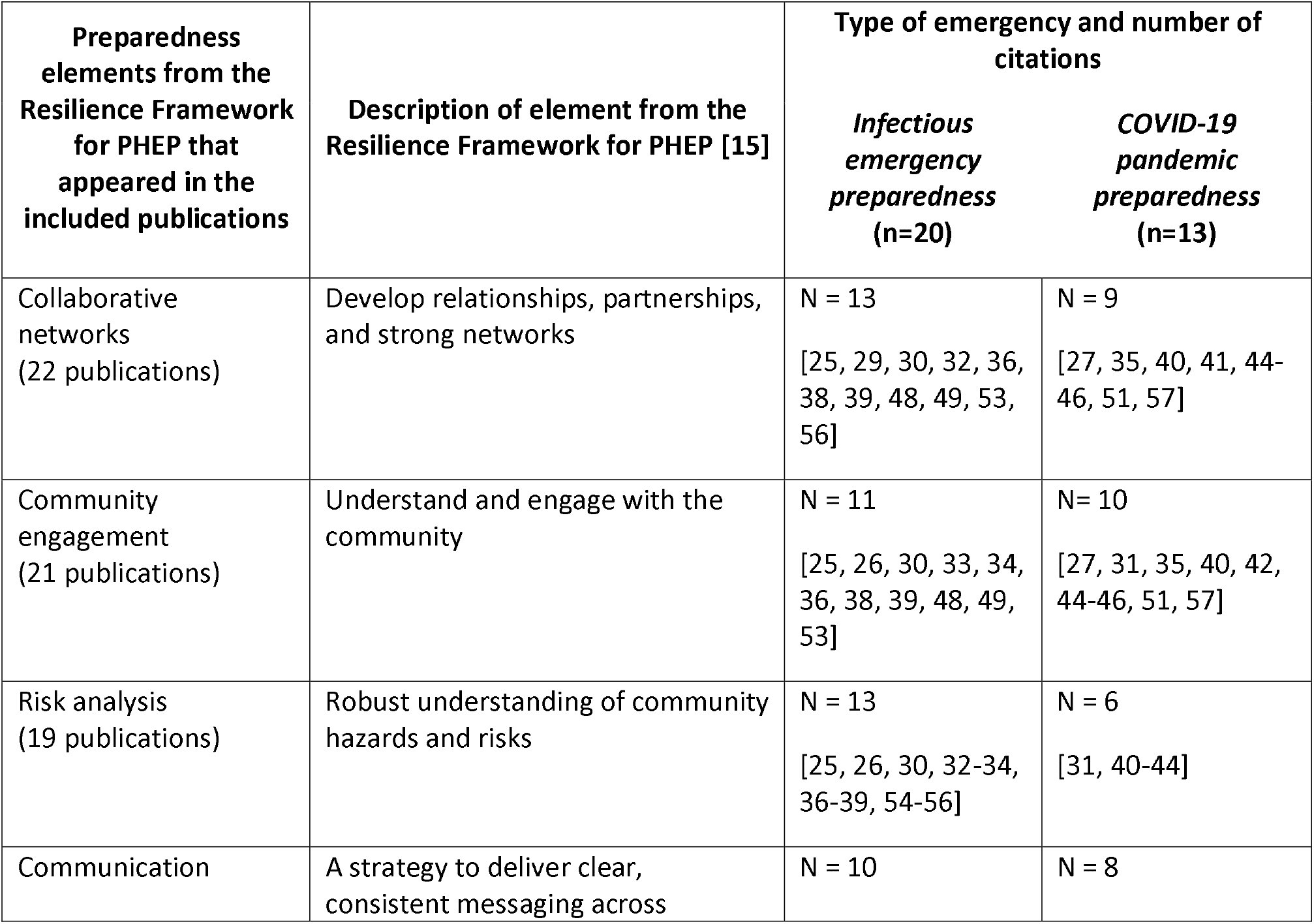
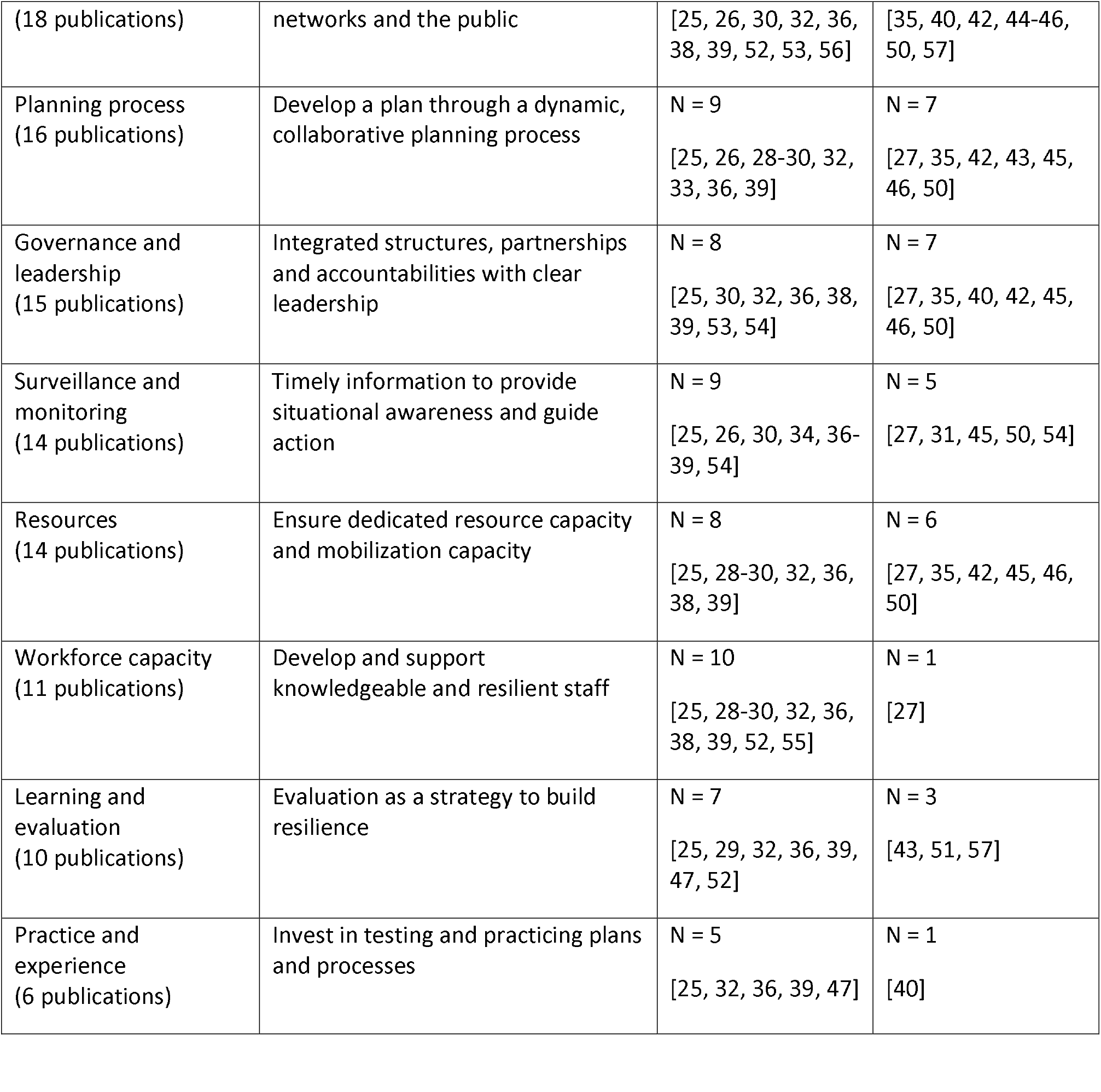
Publications according to emergency type and corresponding Resilience Framework for PHEP elements

### 3.3 Emerging preparedness themes that expand on the elements in the Resilience Framework for PHEP

After comparing and contrasting as part of analysis, our synthesis resulted in the identification of ten themes that expand on the elements in the Resilience Framework for PHEP [15], all with a focus on infectious disease emergency preparedness (see Table 2). Five themes that expand on the framework were observed across both the indexed and grey literature; ordered from most to least common according to the number of publications in which they appear, these themes were: mitigating inequities, vaccination, research and evidence-informed decision making, laboratory and diagnostic system capacity, and infection prevention and control (IPAC) capacity. There were three themes that expand on the Resilience Framework for PHEP that emerged solely from the indexed literature (climate and environmental health, public health legislation, phases of preparedness) and two that emerged solely from the grey literature (financial investment in infrastructure, and health system capacity).

**Table 2.** Publications according to emergency type and emergent themes expanding on the Resilience Framework for PHEP.

| Emergent themes from the included publications | Description of emergent theme | Type of emergency and number of citations |  |
| --- | --- | --- | --- |
|  |  | *Infectious emergency preparedness* (n=18) | *COVID-19 pandemic preparedness* (n=11) |
| Mitigating inequities (19 publications) | The anticipation and mitigation of inequitable impacts of emergencies and public health measures implemented on marginalized, racialized, or other high-risk populations. | N = 10<br>[25, 26, 33, 34, 36-39, 49, 53] | N = 9<br>[31, 35, 40-45, 51] |
| Research and evidence-informed decision making (8 publications) | Building capacity for knowledge-sharing networks and the integration of data-, scientific- and evidence-informed decision-making when preparing and planning for infectious disease emergencies. | N = 5<br>[29, 39, 53-55] | N = 3<br>[27, 40, 51] |
| Vaccination (7 publications) | Preparation for vaccine research, procurement, distribution, education, prioritization of administration to population groups, and any processes related to vaccine policies. | N = 2<br>[30, 54] | N = 5<br>[40, 44-46, 51] |
| Laboratory and diagnostic system capacity (5 publications) | Expanded and clearly defined roles for laboratory and diagnostic systems in infectious disease preparedness plans. | N = 2<br>[30, 54] | N = 3<br>[27, 50, 51] |
| Infection prevention and control (IPAC) capacity (5 publications) | Expanded and clearly defined roles for IPAC capacities, supplies and education. | N = 4<br>[32, 36, 38, 52] | N = 1<br>[51] |
| Financial investment in infrastructure (3 publications) | Adequate preparedness capacities require financial resources to establish critical infrastructure, including sustainable commitment and funding. | N = 2<br>[53, 54] | N = 1<br>[57] |
| Health system capacity | Health system planning should consider the system's surge capacity to safely and effectively care for patients during | N = 2 | N = 1 |
|  |  | *Infectious emergency preparedness* (n=18) | *COVID-19 pandemic preparedness* (n=11) |
| (3 publications) | an infectious disease emergency, the capacity to maintain essential health services, as well as determining monitoring mechanisms to assess the capacity to continue delivering essential health services throughout the pandemic. | [53, 55] | [51] |
| Climate and environmental health considerations (3 publications) | Consider expanded and defined roles for climate and environmental health expertise in PHEP (i.e., One Health, such as considering the impact of environmental degradation on risk of zoonotic disease with pandemic potential; impact of waste generated by pandemic response operations on the environment). | N = 2<br>[38, 39] | N = 1<br>[40] |
| Public health legislation (3 publications) | Understand the scope, limitations and implications of public health laws, policies and authorities of the region (e.g., emergency use authorization), and how these may interface with other authorities. | N = 3<br>[25, 38, 39] | N = 0 |
| Phases of preparedness (2 publications) | Delineating operational phases within the preparedness component of the cycle may support the organization and operationalization of preparedness to prioritize and implement PHEP activities. | N = 2<br>[32, 39] | N = 0 |

Most publications described activities that should take place while planning or preparing for infectious disease emergencies to operationalize priority areas of preparedness [25-29, 32, 33, 35, 36, 38-40, 42-46, 48, 49, 51, 52, 55, 57]. Activities correspond with various preparedness priority areas and exemplify actions that would be taken during infectious disease emergency preparedness processes. These activities were described in publications in addition to or in place of indicators. Activities were described in a variety of ways across publications, and included steps, actions, suggestions, outcomes or outputs of infectious emergency preparedness planning processes.

Multiple studies from the indexed literature described activities related to the operationalization of preparedness [25-29, 32, 33, 35, 36, 38-40, 42-46, 48, 49]. For example, Jesus et al. (2021)’s model for disability-inclusiveness in pandemic preparedness provided several preparedness activities, some of which included developing intersectoral disability-inclusive pandemic preparedness, using evidence on how to reduce disability disparities to inform planning, and the reinforcement of disability-rights in health professionals’ education [33]. AuYoung et al. (2022) developed general strategies for COVID-19 vaccine hesitancy among marginalized communities relevant to future public health emergencies [44]. Examples of AuYoung et al.’s strategies include increasing community and academic capacity to enhance community-academic partnerships, investing in trusted messengers, increasing the trustworthiness of academic institutions and developing long-term cross-site partnerships [44]. Tan et al. (2021) investigated qualitative factors related to pandemic preparedness and identified strategies to achieve a more holistic and equitable approach to preparedness [40]. According to Tan et al., the ongoing translation of changing scientific evidence into policy actions and the development of trusted communication through effective knowledge translation practices are essential strategies to achieve evidence-informed decision-making in pandemic preparedness [40]. Tan et al. also put forward suggestions related to ecological determinants of health which overlap with disaster risk reduction strategies [61, 62], including addressing the effect of health services on the environment, recognizing the impact of climate and environmental degradation on risk of zoonotic disease, and setting climate goals [40].

Several preparedness frameworks identified in the grey literature included preparedness activities, outputs or outcomes [51, 52, 55, 57]. The *WHO’s Strategic Preparedness, Readiness and Response Plan to End the Global COVID-19 Emergency in 2022* describes approaches to managing misinformation, such as peer-to-peer interventions to help communities identify accurate vaccine information by building resilience against misinformation [51]. The WHO’s *Strategic Toolkit for Assessing Risks for All-hazards Emergencies* lists expected activities and outputs of applying the toolkit’s six steps, one such activity is a gap analysis that can inform health and public health workforce capacity building [55]. The WHO’s *Risk Communication and Community Engagement* tool included a list of open-ended questions intended for use within focus group discussions or key informant interviews to support preparedness planning for risk communication and community engagement [57]. Together, these activities and outputs help to operationalize priority areas of infectious disease preparedness.

### 3.4 Preparedness Indicators

In the final step of analysis we examined studies for available indicators or actions/activities to inform indicator development. Compared to the literature identified on frameworks and priority areas for preparedness, there were comparatively fewer indexed and grey literature records identified that describe qualitative and quantitative preparedness indicators. Five indexed studies [30, 31, 34, 37, 41] and three grey literature documents [58-60] either included or focused on describing indicators for pandemic and infectious disease preparedness. It is worth noting that the quantitative indicators identified in the publications (i.e., budget, vaccination targets) largely did not provide specific quantitative thresholds, allowing them to be tailored to various public health agencies’ contexts (e.g., local, regional or provincial).

The types of infectious disease preparedness indicators identified in this scoping review measured or assessed various areas of preparedness including the equity impacts of emergencies [31, 41, 59], core public health and government capacities for emergency preparedness and response [30, 60], population and healthcare system vulnerabilities during pandemics [37], community readiness [34], and benchmarks to strengthen health systems during outbreaks [58]. Some examples of indicators related to public health and health system readiness or capacity include: adequate public health budget [59, 60], capacity to deliver vaccines and the proportion of the population getting vaccinated [30, 58, 60], licensed nurses’ ability to practice in other regions or states [60], oversight of research on dangerous pathogens [58], and enhanced training for the safe transportation of biohazards [58].

Some examples of equity-related preparedness indicators identified through this review are: proportion of population in a defined region who are racialized or first-generation immigrants [31], benchmarks for public health agency plans to embed the needs of racialized or marginalized populations [59], proportion of population with access to internet and technology [41], ratio of residential and nursing homes per 10,000 population aged over 70 years old [37], proportion of population with access to clean water [60], and the proportion of households with at least one of the following: no kitchen, no plumbing, high cost of living, or overcrowded living conditions [34].

## 4.0 Discussion

This scoping review examined the recent literature on conceptualizing, defining and measuring PHEP, which is of relevance to public health agencies as they continue to respond to the COVID-19 pandemic, and remain ready for future infectious disease emergencies. Recent literature on infectious disease emergencies was compared with an evidence-based Resilience Framework for PHEP, which encompasses both infectious and non-infectious emergencies [15, 16]. In general, there was alignment between the themes that emerged from the studies identified and the elements in the Resilience Framework for PHEP. In particular, collaborative networks, community engagement, risk analysis and communication were the framework elements frequently observed across the publications included in this review. This review also revealed emergent themes that expand on the Resilience Framework for PHEP, including mitigating inequities, scientific capacity (research and evidence-informed decision making) and public health capacity (vaccination, laboratory capacity, IPAC capacity). These emergent themes represent areas of PHEP that warrant enhanced consideration for infectious disease emergency preparedness.

Mitigating inequities emerged as an important theme across many included publications. Population health inequities were present and known prior to the COVID-19 pandemic; however, the pandemic re-focused attention on the need for equity-oriented actions due to the inequitable burden of COVID-19 morbidity and mortality, and disproportionate impact of both implementation and removal of pandemic-related response measures among marginalized communities [7-9]. Several publications highlighted the importance of anticipating and mitigating inequitable impacts resulting from infectious disease emergencies and related emergency response measures on marginalized populations. Studies described a variety of equity considerations for preparedness, including the importance of monitoring baseline population characteristics, fostering community trust, and planning for material or financial supports for those inequitably impacted. In addition, infectious disease preparedness frameworks identified in the grey literature provided examples of preparedness activities that help to mitigate inequities related to infectious public health emergencies, including the engagement of trusted community members to ensure communications reach marginalized populations [63]. The Resilience Framework for PHEP includes ethics and values as a concept that is core to all elements in the framework, and included some indicators within the elements to support ethics-informed preparedness actions for local/regional public health agencies. The emergent theme of mitigating inequities in this scoping review reinforces that equity should also be explicitly incorporated as a foundational component of future preparedness frameworks, efforts and actions.

Research and evidence-informed decision-making are central concepts in public health practice and important for emergency preparedness. This theme was often discussed in the indexed literature and was the most frequently observed theme in the grey literature. These publications discussed the importance of knowledge-sharing networks, building capacities for data collection, analysis, and research generation to ensure that infectious disease preparedness activities are evidence-informed. This emergent theme is an example of how themes identified through this scoping review intersect and overlap with other emergent themes as well as elements of the Resilience Framework for PHEP [15, 16]. For example, building capacity for research and evidence-informed decision-making across governments, communities and non-governmental agencies requires action related to mitigating inequities, communication, community engagement, collaborative networks, surveillance and monitoring, among others.

Vaccination, laboratory capacity, climate health, and public health legislation are additional preparedness considerations that reflect changes to PHEP planning that were in progress before the pandemic and have received renewed attention during the COVID-19 pandemic. Public confidence in vaccination is an example of the need for management of public health misinformation that pre-dates, and was further exacerbated by, the COVID-19 pandemic. Several publications discussed vaccination and laboratory systems as key components of the COVID-19 response as well as important priority areas for future pandemic planning. Considerations for climate and environmental health and public health legislation are broad topics that have garnered renewed attention for preparedness in light of the COVID-19 pandemic. Publications that included climate or environmental considerations noted the complex relationship between these issues and potential future pandemics due to climate change and environmental degradation increasing the risk of zoonotic diseases crossing over to humans, and the contributions of healthcare and pandemic response operations to waste, emissions and potential environmental contamination. While governance and leadership is a cross-cutting element in the Resilience Framework for PHEP, the COVID-19 pandemic has renewed an interest in clearly articulating public health emergency roles and responsibilities in public health legislation; thus, providing legislative or policy support for public health emergency decision-making [64].

Themes that expand on the Resilience Framework for PHEP [15, 16] represent potential areas for improvement in the public health and the health system in general, and are not all specific to infectious disease and pandemic preparedness. For example, the domains of financial investment in infrastructure, and health system capacity are areas of focus with population-level health benefits that extend beyond preparedness for public health emergencies and infectious diseases. Several publications highlighted the need for investments to build strong and resilient health and public systems to mitigate the impacts of a health emergency and reduce disruption to essential health and public health services. The COVID-19 pandemic may have exposed these areas of weakness; however, these aspects of the public health system required attention and improvement prior to and beyond the pandemic as noted in previous reports on the impact of the SARS-CoV-1 pandemic [65, 66]. While pandemic and emergency-specific surge capacities and plans are needed, an adequate and resilient baseline is also required.

This scoping review identified emerging priority areas for action in infectious disease emergency preparedness; however, there were comparatively fewer records identified that describe qualitative and quantitative preparedness indicators published since evidence-based indicator development by Khan et al [17]. Detailed analysis, evaluation and indicator development was beyond the scope of this review, although the findings suggest areas of focus that should be considered in future planning. Of note, an exploratory analysis of pandemic preparedness compared with pandemic outcomes posited that some existing preparedness indices are not well suited to predicting pandemic outcomes, but instead are better served as tools to highlight gaps in pandemic capacities [67]. Further work is needed in development of indicators, and also their validation in relation to relevant outcomes. In addition, continued work is needed to ensure preparedness is reinforced as a dynamic, adaptive concept, consistent with complex adaptive systems theory. Anchoring preparedness, planning, and readiness as upstream activities to support resilience of the system can support the concept of the work as continuous improvement and adaptation [16]. The COVID-19 pandemic continues to evolve globally, and jurisdictions are still engaged in the pandemic response and may not yet have capacity to explore how the pandemic might change their approach to emergency planning moving forward. In the coming months and years, there will likely be additional evidence to shape future preparedness planning that will include elements and/or indicators that will support effective infectious disease emergency preparedness. This future work should be revisited, examined and documented to ensure that learning from the pandemic response is included in future preparedness planning domains, activities and indicators.

### 4.1 Limitations

A limitation of this scoping review, common to review methodology, is that some relevant records may not have been included. Further, any ongoing work in jurisdictions and academia to update preparedness plans may not be publicly available since the pandemic response is ongoing, or may not be available in English. Although the search strategy employed was detailed and developed by library informational specialists, any key terms not included in the search may have led to some documents being excluded from our findings.

## 5.0 Conclusion

The 11 elements of an evidence-based pre-COVID-19 pandemic, all-hazards Resilience Framework for PHEP developed in Canada relevant to local or regional public health agencies continue to be reflected in the literature identified in this scoping review. In the studies identified in this review, the following elements in the Resilience Framework for PHEP were represented in descending order of frequency: collaborative networks, community engagement, risk analysis, communication, planning process, governance and leadership, surveillance and monitoring, resources, workforce capacity, learning and evaluation, and practice and experience. With the recent global experience of the COVID-19 pandemic, a focus for local/regional public health agency preparedness oriented towards the 11 elements in the Resilience Framework for PHEP, with enhancements for infectious disease preparedness noted in this synthesis, is supported by evidence. This scoping review focused on infectious disease emergencies and through our analysis identified areas for action for ongoing preparedness that pertain to the themes of mitigating inequities, public health capacities, scientific capacities, and considerations for health system capacity. Future work can advance knowledge related to the areas for action identified, into evidence-informed indicators. Strategies and indicators for mitigating inequities should be considered an urgent focus for action to support the reduction of health inequities anticipated for future emergencies.

## Supporting information

Supplemental-search terms

## Data Availability

All data produced in the present work are contained in the manuscript

## List of abbreviations

COVID-19: Coronavirus Disease 2019
IPAC: Infection Prevention and Control
OECD: Organization for Economic Co-operation and Development
PHEP: Public health emergency preparedness
PHO: Public Health Ontario
SARS: Severe acute respiratory syndrome
WHO: World Health Organization

## Declarations

### Ethics approval and consent to participate

Not applicable.

### Consent for publication

Not applicable.

### Availability of data and materials

Not applicable.

## Authors’ contributions

YK, JML, RJ, KES, FG, SKO and MM conceptualized the scoping review. JML, RJ, KES and FG identified, selected, extracted data in the included studies. JML, RJ, KES and FG wrote the manuscript of the scoping review with critical inputs and appraisal from YK and SKO. The manuscript was then reviewed by LEB, BS, MPL, MM and TLOS. All authors have read and approved the manuscript.

## Acknowledgements

The authors gratefully acknowledge the support of Public Health Ontario’s Library Information Specialists, for their contributions to the indexed database and grey literature search strategies. The authors also acknowledge reviewers of earlier versions of this work, including Lori McKinnon of Public Health Ontario.

## Figures, tables and additional files

**Figure 2.** [submitted in separate file]

Title: Flow chart of included records from indexed databases and grey literature searches

File format: Microsoft Word (.docx)

**Additional File 1** [submitted in separate file]

Title: Indexed database and grey literature search queries

File format: Microsoft Word (.docx)

Description: Details of the search strategies employed in the indexed literature and grey literature.

## References

1. Stoddard M, Sarkar S, Yuan L, Nolan RP, White DE, White LF, et al. Beyond the new normal: assessing the feasibility of vaccine-based suppression of SARS-CoV-2. PLOS One. 2021;16(7):e0254734.

2. Our World in Data. Cumulative confirmed COVID-19 deaths. Global Change Data Lab, Oxford. 2022. https://ourworldindata.org/explorers/coronavirus-data-explorer?zoomToSelection=true&time=2020-03-01..latest&facet=none&pickerSort=desc&pickerMetric=total_cases&hideControls=true&Metric=Confirmed+deaths&Interval=Cumulative&Relative+to+Population=false&Color+by+test+positivity=false&country=~OWID_WRL. Accessed 11 Oct 2022.

3. Fussell E, Sastry N, Vanlandingham M. Race, socioeconomic status, and return migration to New Orleans after Hurricane Katrina. Popul Environ. 2010;31(1-3):20–42.

4. Tricco AC LE, Soobiah C, Perrier L, Straus SE. Impact of H1N1 on socially disadvantaged populations: systematic review. PLoS ONE. 2012;7(6):e39437.

5. Wellesley Institute. Tracking COVID-19 through race-based data. Wellesley Institute, Toronto. 2021. https://www.wellesleyinstitute.com/wp-content/uploads/2021/08/Tracking-COVID-19-Through-Race-Based-Data_eng.pdf. Accessed 16 May 2022.

6. Khanijahani A, Iezadi S, Gholipour K, Azami-Aghdash S, Naghibi D. A systematic review of racial/ethnic and socioeconomic disparities in COVID-19. Int J Equity Health. 2021;20(1):248.

7. Public Health Agency of Canada; Tam T. From risk to resilience: an equity approach to COVID-19. Public Health Agency of Canada, Ottawa. 2020. https://www.canada.ca/en/public-health/corporate/publications/chief-public-health-officer-reports-state-public-health-canada/from-risk-resilience-equity-approach-covid-19.html. Accessed 15 Aug 2022.

8. Ismail SJ, Tunis MC, Zhao L, Quach C. Navigating inequities: a roadmap out of the pandemic. BMJ Global Health. 2021;6(1):e004087.

9. McGrail K, Morgan J, Siddiqi A. Looking back and moving forward: addressing health inequities after COVID-19. Lancet Reg Health Am. 2022;9:100232.

10. Sabin NS, Calliope AS, Simpson SV, Arima H, Ito H, Nishimura T, et al. Implications of human activities for (re)emerging infectious diseases, including COVID-19. J Physiol Anthropol. 2020;39(1):29.

11. Venkatesan P. Re-emergence of infectious diseases associated with the past. Lancet Microbe. 2021;2(4):e140.

12. McCloskey B, Dar O, Zumla A, Heymann DL. Emerging infectious diseases and pandemic potential: status quo and reducing risk of global spread. Lancet Infect Dis. 2014;14(10):1001–10.

13. Khan Y, Fazli G, Henry B, de Villa E, Tsamis C, Grant M, et al. The evidence base of primary research in public health emergency preparedness: a scoping review and stakeholder consultation. BMC Public Health. 2015;15:432.

14. World Health Organization. A strategic framework for emergency preparedness. World Health Organization, Geneva. 2017. https://extranet.who.int/sph/sites/default/files/document-library/document/Preparedness-9789241511827-eng.pdf. Accessed 27 Apr 2022.

15. Ontario Agency for Health Protection and Promotion (Public Health Ontario). Public health emergency preparedness framework and indicators: a workbook to support public health practice. King’s Printer for Ontario, Toronto. 2020. https://www.publichealthontario.ca/-/media/Documents/W/2020/workbook-emergency-preparedness.pdf?sc_lang=en. Accessed 27 Apr 2022.

16. Khan Y, O’Sullivan T, Brown A, Tracey S, Gibson J, Généreux M, et al. Public health emergency preparedness: a framework to promote resilience. BMC Public Health. 2018;18(1):1344.

17. Khan Y, Brown AD, Gagliardi AR, O’Sullivan T, Lacarte S, Henry B, et al. Are we prepared? The development of performance indicators for public health emergency preparedness using a modified Delphi approach. PLoS ONE. 2019;14(12):e0226489.

18. Arksey H, O’Malley L. Scoping studies: towards a methodological framework. Int J Soc Res Methodol. 2005;8(1):19–32.

19. Peters MD, Godfrey CM, Khalil H, McInerney P, Parker D, Soares CB. Guidance for conducting systematic scoping reviews. Int J Evid Based Healthc. 2015 Sep;13(3):141–6.

20. Levac D, Colquhoun H, O’Brien KK. Scoping studies: advancing the methodology. Implement Sci. 2010;5(1):69.

21. Khan Y, Sanford S, Sider D, Moore K, Garber G, de Villa E, et al. Effective communication of public health guidance to emergency department clinicians in the setting of emerging incidents: a qualitative study and framework. BMC Health Serv Res. 2017;17(1):312.

22. Nelson C, Lurie N, Wasserman J, Zakowski S. Conceptualizing and defining public health emergency preparedness. Am J Public Health. 2007;97 Suppl 1(Suppl 1):S9–11.

23. Paek HJ, Hilyard K, Freimuth V, Barge JK, Mindlin M. Theory-based approaches to understanding public emergency preparedness: implications for effective health and risk communication. J Health Commun. 2010;15(4):428–44.

24. United Kingdom. National Health Service Institute for Innovation and Improvement. The good indicators guide: understanding how to use and choose indicators. National Health Service, London. 2017. https://www.england.nhs.uk/improvement-hub/wp-content/uploads/sites/44/2017/11/The-Good-Indicators-Guide.pdf. Accessed 27 Apr 2022.

25. Savoia E, Lin L, Bernard D, Klein N, James LP, Guicciardi S. Public health system research in public health emergency preparedness in the United States (2009–2015): actionable knowledge base. Am J Public Health. 2017;107(S2):e1–e6.

26. Williams BE, Kondo KK. Preventing unequal health outcomes in COVID-19: a systematic review of past interventions. 2021;5(1):856–71.

27. Yoon YK, Lee J, Kim SI, Peck KR. A systematic narrative review of comprehensive preparedness strategies of healthcare resources for a large resurgence of COVID-19 nationally, with local or regional epidemics: present era and beyond. J Korean Med Sci. 2020;35(44).

28. Aruru M, Truong HA, Clark S. Pharmacy emergency preparedness and response (PEPR): a proposed framework for expanding pharmacy professionals’ roles and contributions to emergency preparedness and response during the COVID-19 pandemic and beyond. Res Social Adm Pharm. 2021;17(1):1967–77.

29. Bardosh KL, de Vries DH, Abramowitz S, Thorlie A, Cremers L, Kinsman J, et al. Integrating the social sciences in epidemic preparedness and response: a strategic framework to strengthen capacities and improve global health security. Global Health. 2020;16(1):120.

30. Boyce MR, Katz R. Rapid urban health security assessment tool: a new resource for evaluating local-level public health preparedness. BMJ Glob Health. 2020;5(6):e002606.

31. Brakefield WS, Ammar N, Olusanya OA, Shaban-Nejad A. An urban population health observatory system to support COVID-19 pandemic preparedness, response, and management: design and development study. JMIR Public Health Surveill. 2021;7(6):e28269.

32. de Rooij D, Belfroid E, Eilers R, Roßkamp D, Swaan C, Timen A. Qualitative research: institutional preparedness during threats of infectious disease outbreaks. Biomed Res Int. 2020; doi:10.1155/2020/5861894.

33. Jesus TS, Kamalakannan S, Bhattacharjya S, Bogdanova Y, Arango-Lasprilla JC, Bentley J, et al. PREparedness, REsponse and SySTemic transformation (PRE-RE-SyST): a model for disability-inclusive pandemic responses and systemic disparities reduction derived from a scoping review and thematic analysis. Int J Equity Health. 2021;20(1):204.

34. Links JM, Schwartz BS, Lin S, Kanarek N, Mitrani-Reiser J, Sell TK, et al. COPEWELL: a conceptual framework and system dynamics model for predicting community functioning and resilience after disasters. Disaster Med Public Health Prep. 2018;12(1):127–37.

35. Maqbool A, Khan NZ. Analyzing barriers for implementation of public health and social measures to prevent the transmission of COVID-19 disease using DEMATEL method. Diabetes Metab Syndr. 2020;14(5):887–92.

36. Meyer D, Bishai D, Ravi SJ, Rashid H, Mahmood SS, Toner E, et al. A checklist to improve health system resilience to infectious disease outbreaks and natural hazards. BMJ Glob Health. 2020;5(8):e002429.

37. Nicodemo C, Barzin S, Cavalli N, Lasserson D, Moscone F, Redding S, et al. Measuring geographical disparities in England at the time of COVID-19: results using a composite indicator of population vulnerability. BMJ Open. 2020;10(9):e039749.

38. Sell TK, Shearer MP, Meyer D, Chandler H, Schoch-Spana M, Thomas E, et al. Public health resilience checklist for high-consequence infectious diseases-informed by the domestic ebola response in the United States. J Public Health Manag Pract. 2018;24(6):510–8.

39. Tagarev T, Ratchev V. A taxonomy of crisis management functions. Sustainability. 2020;12(12):5147.

40. Tan MMJ, Neill R, Haldane V, Jung A-S, De Foo C, Tan SM, et al. Assessing the role of qualitative factors in pandemic responses. BMJ. 2021;375:e067512.

41. Wong EY, Schachter A, Collins HN, Song L, Ta ML, Dawadi S, et al. Cross-sector monitoring and evaluation framework: social, economic, and health conditions impacted during the COVID-19 pandemic. Am J Public Health. 2021;111(S3):S215–S23.

42. Choi H, Kim S-Y, Kim J-W, Park Y, Kim M-H. Mainstreaming of health equity in infectious disease control policy during the COVID-19 pandemic era. J Prev Med Public Health. 2021;54(1):1–7.

43. Glover RE, van Schalkwyk MCI, Akl EA, Kristjannson E, Lotfi T, Petkovic J, et al. A framework for identifying and mitigating the equity harms of COVID-19 policy interventions. J Clin Epidemiol. 2020;128:35–48.

44. AuYoung M, Rodriguez Espinosa P, Chen W-T, Juturu P, Young M-EDT, Casillas A, et al. Addressing racial/ethnic inequities in vaccine hesitancy and uptake: lessons learned from the California alliance against COVID-19. J Behav Med. 2022.

45. Blouin Genest G, Burlone N, Champagne E, Eastin C, Ogaranko C. Translating COVID-19 emergency plans into policy: a comparative analysis of three Canadian provinces. Policy Des Pract. 2021;4(1):115–32.

46. Schulze C, Welker A, Kühn A, Schwertz R, Otto B, Moraldo L, et al. Public health leadership in a VUCA world environment: lessons learned during the regional emergency rollout of SARS-CoV-2 vaccinations in Heidelberg, Germany, during the COVID-19 pandemic. Vaccines. 2021;9(8).

47. Reddin K, Bang H, Miles L. Evaluating simulations as preparation for health crises like CoVID-19: insights on incorporating simulation exercises for effective response. Int J Disaster Risk Reduct. 2021;59:102245.

48. Schoch-Spana M, Ravi S, Meyer D, Biesiadecki L, Mwaungulu GJ. High-performing local health departments relate their experiences at community engagement in emergency preparedness. J Public Health Manag Pract. 2018;24(4):360–9.

49. Schoch-Spana M, Nuzzo J, Ravi S, Biesiadecki L, Mwaungulu G, Jr. The local health department mandate and capacity for community engagement in emergency preparedness: a national view over time. J Public Health Manag Pract. 2018;24(4):350–9.

50. Lee CT, Buissonnière M, McClelland A, Frieden TR. Association between preparedness and response measures and COVID-19 incidence and mortality. medRxiv. 2021. doi:10.1101/2021.02.02.21251013.

51. World Health Organization. Strategic preparedness, readiness and response plan to end the global COVID-19 emergency in 2022. World Health Organzation, Geneva. 2022. https://www.who.int/publications/i/item/WHO-WHE-SPP-2022.1. Accessed 27 Apr 2022.

52. World Health Organization. Framework and toolkit for Infection prevention and control in outbreak preparedness, readiness and response at the national level. World Health Organization, Geneva. 2021. https://apps.who.int/iris/bitstream/handle/10665/345251/9789240032729-eng.pdf?sequence=1&isAllowed=y. Accessed 27 Apr 2022.

53. World Health Organization. Framework for strengthening health emergency preparedness in cities and urban settings. World Health Organization, Geneva. 2021. https://www.who.int/publications/i/item/9789240037830. Accessed 27 Apr 2022

54. World Health Organization. Pandemic influenza preparedness (PIP) framework for the sharing of influenza viruses and access to vaccines and other benefits, second edition. World Health Organization, Geneva. 2022. https://www.who.int/publications/i/item/9789240024854. Accessed 27 Apr 2022.

55. World Health Organization. Strategic toolkit for assessing risks: a comprehensive toolkit for all-hazards health emergency risk assessment. World Health Organization, Geneva. 2021. https://www.who.int/publications/i/item/9789240036086. Accessed 27 Apr 2022.

56. World Health Organization, Food and Agricultre Organization of the United Nations. Joint risk assessment operational tool (JRA OT): an operational tool of the tripartite zoonoses guide. World Health Organization, Geneva. 2020. https://apps.who.int/iris/bitstream/handle/10665/340005/9789240015142-eng.pdf?sequence=1&isAllowed=y. Accessed 27 Apr 2022

57. World Health Organization. Risk communication and community engagement action plan guidance: COVID-19 preparedness & response. World Health Organization, Geneva. 2020. https://www.who.int/publications/i/item/risk-communication-and-community-engagement-(rcce)-action-plan-guidance. Accessed 27 Apr 2022.

58. Bell JA, Nuzzo JB, Britsol N, Essix G, Isaac C, Kobokovich A, et al. Global health security index 2021: advancing collective action and accountability amid global crisis. John’s Hopkins Center for Health Security, Baltimore. 2021. https://www.centerforhealthsecurity.org/our-work/publications/GHSindex2021. Accessed 18 July 2022.

59. National Collaborating Centre for Infectious Diseases. Measuring what counts in the midst of the COVID-19 pandemic: equity indicators for public health [Internet]. National Collaborating Centre for Infectious Diseases, Winnipeg. 2021. https://nccid.ca/publications/measuring-what-counts-in-the-midst-of-the-covid-19-pandemic-equity-indicators-for-public-health/. Accessed 18 Jul 2022.

60. Trust For America’s Health. Ready or not 2022: protecting the public’s health from diseases, disasters, and bioterrorism. Trust for America’s Health, Washington. 2022. https://www.tfah.org/report-details/ready-or-not-2022/. Accessed 18 July 2022.

61. United Nations Office for Disaster Risk Reduction. Sendai framework for disaster risk reduction 2015-2030. United Nations Office for Disaster Risk Reduction, Geneva. 2015. https://www.undrr.org/publication/sendai-framework-disaster-risk-reduction-2015-2030. Accessed 30 Sep 2022.

62. United Nations Framework Convention on Climate Change. Paris Agreement. United Nations Framework Convention on Climate Change, Geneva. 2015. https://unfccc.int/process-and-meetings/the-paris-agreement/the-paris-agreement#:~:text=The%20Paris%20Agreement%20is%20a,compared%20to%20pre%2Dindustrial%20levels. Accessed 03 Sep 2022.

63. World Health Organization. Commission on social determinants of health final report. World Health Organization, Geneva. 2008. https://www.who.int/publications/i/item/9789241563703. Accessed 06 Sept 2022.

64. Network for Public Health Law and National Association of County and City Health Officials. Proposed limits on public health authority: dangerous for public health. Network for Public Health Law, Edina. 2021. https://www.naccho.org/uploads/downloadable-resources/Proposed-Limits-on-Public-Health-Authority-Dangerous-for-Public-Health-FINAL-5.24.21pm.pdf. Accessed 21 Sep 2022.

65. Health Canada. Learning from SARS: renewal of public health in Canada. Her Majesty the Queen in Right of Canada, Ottawa. 2003. https://www.canada.ca/content/dam/phac-aspc/migration/phac-aspc/publicat/sars-sras/pdf/sars-e.pdf. Accessed 06 Sept 2022.

66. Campbell A. The SARS commission: SARS and public health in Ontario. Commission to Investigate the Introduction and Spread of SARS in Ontario, Toronto. 2006. http://www.archives.gov.on.ca/en/e_records/sars/report/index.html. Accessed 06 Sept 2022.

67. COVID-19 National Preparedness Collaborators. Pandemic preparedness and COVID-19: an exploratory analysis of infection and fatality rates, and contextual factors associated with preparedness in 177 countries, from Jan 1, 2020, to Sept 30, 2021. Lancet. 2022 Apr 16;399(10334):1489–1512.

