## Supplemental-search terms for "What is the current state of public health system preparedness for infectious disease emergencies? A scoping review"

**S1. Indexed database and grey literature search queries**

**Table 1. MEDLINE search query**

| # | Searches | Results |
| --- | --- | --- |
| 1 | (((Public Health/ or Public Health Practice/ or Public Health Administration/ or Public Health Nursing/ or Public Health Systems Research/ or United States Public Health Service/ or "Centers for Disease Control and Prevention, U.S."/ or Community Health Nursing/ or Community Health Planning/ or Community Health Services/ or Community Integration/ or Community Medicine/ or Community Networks/ or Community Participation/ or Community-Based Participatory Research/ or Community-Institutional Relations/ or Health Education/ or Health Promotion/ or Population Characteristics/ or Population Dynamics/ or Population Groups/ or Population Health Management/ or Population Health/ or Population/ or Rural Population/ or Social Medicine/ or Sociology, Medical/ or Suburban Population/ or Urban Population/ or Epidemics/ or Pandemics/ or Disease Outbreaks/ or COVID-19/ or SARS-CoV-2/ or Hemorrhagic Fever, Ebola/ or Ebolavirus/ or Severe Acute Respiratory Syndrome/ or SARS Virus/ or Influenza A Virus, H1N1 Subtype/ or Zika Virus Infection/ or Zika Virus/ or Middle East Respiratory Syndrome Coronavirus/ or "SARS-CoV-2 variants".os. or "COVID-19 breakthrough infections".rs.) and ((Emergencies/ not (exp Emergency Service, Hospital/ or Emergency Medical Services/ or exp Surgical Procedures, Operative/)) or Disaster Victims/ or Disasters/ or Mass Casualty Incidents/ or Disaster Planning/ or ((emergency or emergencies or disaster*) not ((emergency or emergencies) adj2 (accident or care or service* or department* or room or rooms or unit or units or ward or wards or hospital* or medical* or surgery or surgeries or surgical* or visit* or doctor* or physician*))).kf,kw,ti.)) not medline.st.) or (*Epidemics/ or *Pandemics/ or *Disease Outbreaks/ or *COVID-19/ or *SARS-CoV-2/ or *Hemorrhagic Fever, Ebola/ or *Ebolavirus/ or *Severe Acute Respiratory Syndrome/ or *SARS Virus/ or *Influenza A Virus, H1N1 Subtype/ or *Zika Virus Infection/ or *Zika Virus/ or *Middle East Respiratory Syndrome Coronavirus/ or "SARS-CoV-2 variants".os. or "COVID-19 breakthrough infections".rs.) | 213558 |
| 2 | ((pandemic* or "public health" or "health authorit*" or "health department*" or "health education" or "health promotion" or "health protection" or "health system*" or "health unit*" or community or communities or county or counties or municipal* or neighborhood or neighborhoods or neighbourhood or neighbourhoods or ((region or regions or regional*) adj3 health) or parish or parishes or population or populations or (outbreak* adj5 (diseas* or COVID* or SARS* or "severe acute respiratory syndrome" or ebola* or H1N1* or zika or MERS or "middle east respiratory syndrome")) or epidemic*) and ((emergency or emergencies or disaster*) not ((emergency or emergencies) adj2 (accident or care or service* or department* or room or rooms or unit or units or ward or wards or hospital* or medical* or surgery or surgeries or surgical* or visit* or doctor* or physician*)))).kf,kw,ti. or ("public health emergency" or "public health emergencies" or ((pandemic* or "public health" or epidemic* or (diseas* adj2 (outbreak* or infect* or emerging or communicable)) or COVID* or SARS* or "severe acute respiratory syndrome" or ebola* or H1N1* or zika or MERS or "middle east respiratory syndrome" or epidemic*) adj5 (emergency or emergencies or disaster*))).ti,kf,kw. | 8086 |
| 3 | 1 or 2 | 219138 |
| 4 | Civil Defense/ or Disaster Medicine/ or Disaster Planning/ or Adaptation, Psychological/ or Resilience, Psychological/ or Posttraumatic Growth, Psychological/ or Relief Work/ or Health Plan Implementation/ or Health Planning Guidelines/ or Guideline Adherence/ or Health Planning/ or Health Systems Plans/ or Regional Health Planning/ or Social Planning/ or State Health Plans/ or Strategic Planning/ or Capacity Building/ or Government Programs/ or Pilot Projects/ or Program Development/ or Government Regulation/ or Health Policy/ or Public Policy/ or Risk Management/ or Forecasting/ or Resource Allocation/ or Safety Management/ or Security Measures/ or International Health Regulations/ or International Cooperation/ or National Health Programs/ or ((prepar* or plan or plans or plann* or manag* or train* or resilien* or coping or cope or respon* or ready or readiness or recover* or rebuild* or "build* back" or "bounc* back" or countermeasure*).ti,kf,kw. not medline.st.) or ("public health preparedness" or "public health emergency preparedness" or "pandemic preparedness" or "pandemic emergency preparedness").ti. | 946944 |
| 5 | COVID-19/pc or Coronavirus Infections/pc or Pandemics/pc or Epidemics/pc or Disease Outbreaks/pc or Pneumonia, Viral/pc or Communicable Diseases, Emerging/pc or Disease Transmission, Infectious/pc or Communicable Disease Control/lj, og or Infection Control/mt, og or Public Health/lj, og or Public Health Practice/lj or Public Health Administration/lj, og or ((((emergencies or emergency) not ((emergency or emergencies) adj2 (accident or care or service* or department* or room or rooms or unit or units or ward or wards or hospital* or medical* or surgery or surgeries or surgical* or visit* or doctor* or physician*))) or pandemic* or epidemic* or disaster* or "mass casualt*" or COVID* or SARS* or "severe acute respiratory syndrome" or future or post-emergenc* or post-pandemic* or post-covid* or post-sars* or (diseas* adj2 (infect* or emerging or communicable))) adj3 (prepar* or plan or plans or plann* or manag* or train* or resilien* or coping or cope or respon* or ready or readiness or recover* or rebuild* or "build* back" or "bounc* back" or countermeasure*)).ti,kw,kf. | 94252 |
| 6 | 4 or 5 | 1016496 |
| 7 | Evaluation Studies as Topic/ or Evaluation Study/ or Evaluation Study.pt. or *Retrospective Studies/ or Systems Analysis/ or Benchmarking/ or Quality Improvement/ or Quality Control/ or Total Quality Management/ or Efficiency, Organizational/ or Efficiency/ or Management Audit/ or Qualitative Research/ or "Task Performance and Analysis"/ or Program Evaluation/ or Delphi Technique/ or "Cost of Illness"/ or Needs Assessment/ or *"Surveys and Questionnaires"/ or Checklist/ or *Disaster Planning/st or Civil Defense/st or Public Health Administration/st or Public Health Practice/st or Public Health/st or Communicable Disease Control/st or Models, Organizational/ or Models, Theoretical/ or Implementation Science/ or *Research Design/ | 992383 |
| 8 | (((evaluat* or apprais* or assess* or benchmark* or improv* or enhanc* or quality or ineffective* or efficac* or effective* or outcome* or audit* or analyz* or analys* or ((preparedness or response) adj3 (capacity or capacities or capability or capabilities)) or (quality adj3 (assess* or improv* or manag* or control* or assur*))) adj3 (reporting or report or measur* or metric* or score* or scoring or scorecard* or standard* or target or targets or "COVID-score*" or "report card*" or perform* or quality or ineffective* or efficac* or effective* or outcome* or lesson* or framework* or indicator* or index or indexes or instrument or instruments or inventories or inventory or registries or registry or implement* or initiativ* or intervention or interventions or model or models or tool or tools or theor* or schema* or pilot* or plan or plans or planning or policy or policies or principles or program or programs or programme or programmes or roadmap* or "road map*" or strategy or strategies)).ti,kf,kw. not medline.st.) or (((lesson* adj2 learn*) or "after-action review*" or "after action review*" or "in-action review*" or "in action review*" or scorecard* or "COVID-score*" or "report card*").ti. not medline.st.) | 217355 |
| 9 | 7 or 8 | 1209736 |
| 10 | 3 and 6 and 9 | 4123 |
| 11 | limit 10 to yr="2018 -Current" | 2553 |
| 12 | 10 and ((201706* or 201707* or 201708* or 201709* or 201710* or 201711* or 201712*).ez. or (201706* or 201707* or 201708* or 201709* or 201710* or 201711* or 201712*).dt. or (201706* or 201707* or 201708* or 201709* or 201710* or 201711* or 201712* or 2017 06* or 2017 07* or 2017 08* or 2017 09* or 2017 10* or 2017 11* or 2017 12* or 2017 jun or 2017 jul or 2017 aug or 2017 sep or 2017 oct or 2017 nov or 2017 dec).dp.) | 99 |
| 13 | 11 or 12 | 2623 |
| 14 | (Afghanistan/ or Argentina/ or Asia, Western/ or Asia/ or Bahrain/ or Bangladesh/ or Belize/ or Bhutan/ or Bolivia/ or Brazil/ or Central America/ or Developing Countries/ or Ecuador/ or El Salvador/ or exp Africa/ or exp Asia, Central/ or exp Asia, Northern/ or exp Asia, Southeastern/ or exp Caribbean Region/ or exp China/ or exp India/ or Far East/ or French Guiana/ or Guatemala/ or Guyana/ or Honduras/ or Iran/ or Iraq/ or Jordan/ or Kuwait/ or Latin America/ or Lebanon/ or Middle East/ or Mongolia/ or Nepal/ or Nicaragua/ or Oman/ or Pakistan/ or Panama/ or Paraguay/ or Peru/ or Qatar/ or Saudi Arabia/ or South America/ or Sri Lanka/ or Suriname/ or Syria/ or Taiwan/ or Turkey/ or United Arab Emirates/ or Uruguay/ or Venezuela/ or Yemen/) not (Austria/ or Baltic States/ or Belgium/ or Chile/ or Colombia/ or Costa Rica/ or Czech Republic/ or Developed Countries/ or Estonia/ or Europe/ or exp Australia/ or exp Canada/ or exp Denmark/ or exp France/ or exp Germany/ or exp Italy/ or exp Japan/ or exp Korea/ or exp Norway/ or exp United Kingdom/ or exp United States/ or Finland/ or Greece/ or Hungary/ or Iceland/ or Ireland/ or Israel/ or Latvia/ or Lithuania/ or Luxembourg/ or Mexico/ or Netherlands/ or New Zealand/ or North America/ or Poland/ or Portugal/ or Slovakia/ or Slovenia/ or Spain/ or Sweden/ or Switzerland/ or Turkey/) | 1114771 |
| 15 | 13 not 14 | 2066 |
| 16 | limit 15 to English | 2034 |
| 17 | remove duplicates from 16 | 2033 |

**Table 2. National Institutes of Health iSearch COVID-19 Portfolio Search Query**

| # | Searches | Results |
| --- | --- | --- |
| 1 | (title:"emergency preparedness"~3 OR title:"emergency plan"~3 OR title:"emergency management"~3 OR title:"emergency response"~3 OR title:"emergency recovery"~3 OR title:"system preparedness"~3 OR title:"system response"~3 OR title:"system recovery"~3 OR title:"community preparedness"~3 OR title:"community response"~3 OR title:"community recovery"~3 OR title:"public-health preparedness"~3 OR title:"public health preparedness"~3 OR title:"public-health response"~3 OR title:"public health response"~3 OR title:"public-health recovery"~3 OR title:"public health recovery"~3 OR title:"public-health plan"~3 OR title:"public health plan"~3 OR title:"pandemic preparedness"~3 OR title:"pandemic response"~3 OR title:"pandemic recovery"~3) AND PubTypes:"Preprint" | 140 |
| 2 | (title:emergenc* OR title:pandemic* OR title:"public health" OR title:public-health OR title:communit* OR title:"health system" OR title:covid* OR title:sars* OR title:coronavirus) AND (title:prepar* OR title:plan OR title:plans OR title:planning OR title:train*) AND (title:evaluat* OR title:assess* OR title:audit* OR title:analyz* OR title:analys* OR title:performance OR title:quality OR title:improve* OR title:enhance* OR title:indicator* OR title:measure* OR title:metric* OR title:scorecard* OR title:score* OR title:scoring OR title:lesson* OR title: lessons-learned OR title:ineffective OR title:efficacy OR title:effective* OR title:outcome* OR title:review* OR title:report*) AND PubTypes:"Preprint" | 69 |
|  | (title:emergenc* OR title:pandemic* OR title:"public health" OR title:public-health OR title:communit* OR title:"health system") AND (title:manag* OR title:respons* OR title:respond* OR title:recover* OR title:resilient* OR title:resilienc* OR title:"build back" OR title:"building back" OR title:"bounce back" OR title:"bouncing back") AND (title:evaluat* OR title:assess* OR title:audit* OR title:analyz* OR title:analys* OR title:performance OR title:quality OR title:improve* OR title:enhance* OR title:indicator* OR title:measure* OR title:metric* OR title:scorecard* OR title:score* OR title:scoring OR title:lesson* OR title:lessons-learned OR title:ineffective OR title:efficacy OR title:effective* OR title:outcome* OR title:review* OR title:report*) AND PubTypes:"Preprint" | 123 |
|  | (title:emergenc* OR title:pandemic* OR title:"public health" OR title:public-health OR title:communit* OR title:"health system" OR title:covid* OR title:sars* OR title:coronavirus) AND (title:prepar* OR title:plan OR title:plans OR title:planning OR title:train*) AND (title:framework* OR title:intervention* OR title:initiative* OR title:model* OR title:pilot* OR title:roadmap* OR title:"road map" OR title:strategy OR title:strategies) AND PubTypes:"Preprint" | 34 |
|  | (title:emergenc* OR title:pandemic* OR title:"public health" OR title:public-health OR title:communit* OR title:"health system") AND (title:manag* OR title:respons* OR title:respond* OR title:recover* OR title:resilient* OR title:resilienc* OR title:rebuild OR title:coping OR title:cope* OR title:"build back" OR title:"building back" OR title:"bounce back" OR title:"bouncing back") AND (title:framework* OR title:intervention* OR title:initiative* OR title:model* OR title:pilot* OR title:roadmap* OR title:"road map" OR title:strategy OR title:strategies) AND PubTypes:"Preprint" | 70 |

**Table 3. Embase search query**

| # | Searches | Results |
| --- | --- | --- |
| 1 | (((public health/ or public health service/ or community health nursing/ or public health systems research/ or health care planning/ or community care/ or community integration/ or community medicine/ or community participation/ or participatory research/ or public relations/ or health education/ or health promotion/ or "population and population related phenomena"/ or population dynamics/ or population group/ or population health management/ or population health/ or population/ or rural population/ or social medicine/ or medical sociology/ or suburban population/ or urban population/ or epidemic/ or pandemic/ or coronavirus disease 2019/ or Severe acute respiratory syndrome coronavirus 2/ or Ebola hemorrhagic fever/ or severe acute respiratory syndrome/ or SARS coronavirus/ or "Influenza A virus (H1N1)"/ or Zika fever/ or Zika virus/ or Middle East respiratory syndrome coronavirus/) and ((emergency/ not (exp hospital emergency service/ or emergency health service/ or exp surgery/)) or disaster victim/ or disaster/ or mass disaster/ or disaster planning/ or ((emergency or emergencies or disaster*) not ((emergency or emergencies) adj2 (accident or care or service* or department* or room or rooms or unit or units or ward or wards or hospital* or medical* or surgery or surgeries or surgical* or visit* or doctor* or physician*))).kf,kw,ti.)) not embase.st.) or (*epidemic/ or *pandemic/ or *coronavirus disease 2019/ or *Severe acute respiratory syndrome coronavirus 2/ or *Ebola hemorrhagic fever/ or *severe acute respiratory syndrome/ or *SARS coronavirus/ or *"Influenza A virus (H1N1)"/ or *Zika fever/ or *Zika virus/ or *Middle East respiratory syndrome coronavirus/) | 253531 |
| 2 | ((pandemic* or "public health" or "health authorit*" or "health department*" or "health education" or "health promotion" or "health protection" or "health system*" or "health unit*" or community or communities or county or counties or municipal* or neighborhood or neighborhoods or neighbourhood or neighbourhoods or ((region or regions or regional*) adj3 health) or parish or parishes or population or populations or (outbreak* adj5 (diseas* or COVID* or SARS* or "severe acute respiratory syndrome" or ebola* or H1N1* or zika or MERS or "middle east respiratory syndrome")) or epidemic*) and ((emergency or emergencies or disaster*) not ((emergency or emergencies) adj2 (accident or care or service* or department* or room or rooms or unit or units or ward or wards or hospital* or medical* or surgery or surgeries or surgical* or visit* or doctor* or physician*)))).kf,kw,ti. or ("public health emergency" or "public health emergencies" or ((pandemic* or "public health" or epidemic* or (diseas* adj2 (outbreak* or infect* or emerging or communicable)) or COVID* or SARS* or "severe acute respiratory syndrome" or ebola* or H1N1* or zika or MERS or "middle east respiratory syndrome" or epidemic*) adj5 (emergency or emergencies or disaster*))).ti,kf,kw. | 8579 |
| 3 | 1 or 2 | 257219 |
| 4 | civil defense/ or disaster medicine/ or disaster planning/ or psychological adjustment/ or psychological resilience/ or "posttraumatic growth (psychology)"/ or relief work/ or health care planning/ or protocol compliance/ or strategic planning/ or capacity building/ or government/ or pilot study/ or program development/ or government regulation/ or health care policy/ or public policy/ or risk management/ or forecasting/ or resource allocation/ or safety/ or "organization and management"/ or international health regulation/ or international cooperation/ or ((prepar* or plan or plans or plann* or manag* or train* or resilien* or coping or cope or respon* or ready or readiness or recover* or rebuild* or "build* back" or "bounc* back" or countermeasure*).ti,kf,kw. not embase.st.) or ("public health preparedness" or "public health emergency preparedness" or "pandemic preparedness" or "pandemic emergency preparedness").ti. | 2157885 |
| 5 | coronavirus disease 2019/pc or Coronavirus infection/pc or pandemic/pc or epidemic/pc or virus pneumonia/pc or communicable disease/pc or disease transmission/pc or communicable disease control/pc or infection control/pc or ((((emergencies or emergency) not ((emergency or emergencies) adj2 (accident or care or service* or department* or room or rooms or unit or units or ward or wards or hospital* or medical* or surgery or surgeries or surgical* or visit* or doctor* or physician*))) or pandemic* or epidemic* or disaster* or "mass casualt*" or COVID* or SARS* or "severe acute respiratory syndrome" or future or post-emergenc* or post-pandemic* or post-covid* or post-sars* or (diseas* adj2 (infect* or emerging or communicable))) adj3 (prepar* or plan or plans or plann* or manag* or train* or resilien* or coping or cope or respon* or ready or readiness or recover* or rebuild* or "build* back" or "bounc* back" or countermeasure*)).ti,kw,kf. | 59191 |
| 6 | 4 or 5 | 2191598 |
| 7 | evaluation study/ or *retrospective study/ or system analysis/ or benchmarking/ or total quality management/ or quality control/ or total quality management/ or organizational efficiency/ or productivity/ or management/ or qualitative research/ or task performance/ or program evaluation/ or program effectiveness/ or program efficacy/ or program impact/ or Delphi study/ or "cost of illness"/ or needs assessment/ or *questionnaire/ or checklist/ or nonbiological model/ or theoretical model/ or implementation science/ or *methodology/ or performance measurement system/ or conceptual framework/ or interview/ or consensus/ or consensus development/ or outcome assessment/ | 1935700 |
| 8 | (((evaluat* or apprais* or assess* or benchmark* or improv* or enhanc* or quality or ineffective* or efficac* or effective* or outcome* or audit* or analyz* or analys* or ((preparedness or response) adj3 (capacity or capacities or capability or capabilities)) or (quality adj3 (assess* or improv* or manag* or control* or assur*))) adj3 (reporting or report or measur* or metric* or score* or scoring or scorecard* or standard* or target or targets or "COVID-score*" or "report card*" or perform* or quality or ineffective* or efficac* or effective* or outcome* or lesson* or framework* or indicator* or index or indexes or instrument or instruments or inventories or inventory or registries or registry or implement* or initiativ* or intervention or interventions or model or models or tool or tools or theor* or schema* or pilot* or plan or plans or planning or policy or policies or principles or program or programs or programme or programmes or roadmap* or "road map*" or strategy or strategies)).ti,kf,kw. not embase.st.) or (((lesson* adj2 learn*) or "after-action review*" or "after action review*" or "in-action review*" or "in action review*" or scorecard* or "COVID-score*" or "report card*").ti. not embase.st.) | 815660 |
| 9 | 7 or 8 | 2617918 |
| 10 | 3 and 6 and 9 | 6486 |
| 11 | limit 10 to yr="2018 -Current" | 4606 |
| 12 | 10 and ((201706* or 201707* or 201708* or 201709* or 201710* or 201711* or 201712*).dd. or (summer 2017 or fall 2017 or autumn 2017 or winter 2017 or jun* 2017 or jul* 2017 or aug* 2017 or sep* 2017 or oct* 2017 or nov* 2017 or dec* 2017).dp.) | 93 |
| 13 | 11 or 12 | 4699 |
| 14 | (Albania/ or Argentina/ or Armenia/ or Aruba/ or Asia/ or Bahrain/ or Balkan Peninsula/ or Belarus/ or Belize/ or Bolivia/ or Bulgaria/ or Central America/ or Croatia/ or Cyprus/ or Eastern Europe/ or Ecuador/ or Egypt/ or El Salvador/ or exp "Bosnia And Herzegovina"/ or exp "Georgia (Republic)"/ or exp Africa/ or exp Azerbaijan/ or exp Brazil/ or exp Caribbean Netherlands/ or exp Caribbean/ or exp Central Asia/ or exp China/ or exp Iraq/ or exp Russian Federation/ or exp Serbia/ or exp South Asia/ or exp Southeast Asia/ or exp Ukraine/ or exp United Arab Emirates/ or Far East/ or French Guiana/ or Guatemala/ or Guyana/ or Honduras/ or Iran/ or Jordan/ or Kosovo/ or Kuwait/ or Lebanon/ or Middle East/ or Moldova/ or Mongolia/ or "Montenegro (Republic)"/ or Netherlands Antilles/ or Nicaragua/ or Northern Asia/ or Oman/ or Palestine/ or Panama/ or Paraguay/ or Peru/ or Philippines/ or Qatar/ or "Republic Of North Macedonia"/ or Romania/ or Saudi Arabia/ or South America/ or Suriname/ or Syrian Arab Republic/ or Taiwan/ or Uruguay/ or Venezuela/ or Western Asia/ or Yemen/) not (Baltic States/ or Chile/ or Colombia/ or Costa Rica/ or Czech Republic/ or Developed Country/ or Estonia/ or Europe/ or exp "Australia And New Zealand"/ or exp Canada/ or exp Mexico/ or exp Southern Europe/ or exp United States/ or exp Western Europe/ or Hungary/ or Israel/ or Japan/ or exp Korea/ or Latvia/ or Lithuania/ or North America/ or Poland/ or Slovakia/ or Slovenia/ or "Turkey (Republic)"/) | 1407730 |
| 15 | 13 not 14 | 3877 |
| 16 | 15 not (editorial or letter or note or conference abstract).pt. | 2782 |
| 17 | limit 16 to english language | 2717 |
| 18 | 17 not (1* or 2* or 3* or 4* or 5* or 6* or 7* or 8* or 9*).pm. | 509 |

**Table 4. Business Continuity and Disaster Recovery Reference Center Search Query**

| # | Query | Limiters/ Expanders | Results |
| --- | --- | --- | --- |
| S1 | ( (DE "PUBLIC health research" OR DE "EPIDEMICS" OR DE "DISEASE outbreaks" OR DE "COMMUNITY health nursing" OR DE "COMMUNITY health services" OR DE "COMMUNITY involvement" OR DE "COMMUNITY-based participatory research" OR DE "HEALTH education" OR DE "HEALTH promotion" OR DE "POPULATION dynamics" OR DE "POPULATION health" OR DE "PUBLIC health administration" OR DE "PUBLIC health nursing" OR DE "PUBLIC health" OR DE "PUBLIC policy (Law)" OR DE "RURAL population" OR DE "SOCIAL medicine" OR DE "CITY dwellers" OR DE "COVID-19" OR DE "COVID-19 pandemic" OR DE "EBOLA virus disease" OR DE "EBOLA virus" OR DE "H1N1 influenza" OR DE "INFLUENZA A virus, H1N1 subtype" OR DE "MERS coronavirus" OR DE "MIDDLE East respiratory syndrome" OR DE "PANDEMICS" OR DE "SARS (Disease)" OR DE "SARS Epidemic, 2002-2003" OR DE "SARS virus" OR DE "SARS-CoV-2" OR DE "ZIKA Virus Epidemic, 2015-" OR DE "ZIKA virus infections" OR DE "ZIKA virus") ) AND ( ( ( DE "EMERGENCIES" NOT ( DE "EMERGENCY medical services" OR DE "AMBULANCE service" OR DE "EMERGENCY medical personnel" OR DE "EMERGENCY services in psychiatric hospitals" OR DE "HOSPITAL emergency services" OR DE "MEDICAL triage" OR DE "PEDIATRIC emergency services" OR DE "POISON control centers" OR DE "SEXUAL assault evidentiary examinations" OR DE "MEDICAL emergencies" OR DE "SURGICAL procedures" OR DE "OPERATIVE surgery" )) ) OR ( TI ( ( (emergency OR emergencies OR disaster*) ) NOT ( (emergency OR emergencies) N2 (accident OR care OR service* OR department* OR room OR rooms OR unit OR units OR ward OR wards OR hospital* OR medical* OR surgery OR surgeries OR surgical* OR visit* OR doctor* OR physician*) ) ) OR KW ( ( (emergency OR emergencies OR disaster*) ) NOT ( (emergency OR emergencies) N2 (accident OR care OR service* OR department* OR room OR rooms OR unit OR units OR ward OR wards OR hospital* OR medical* OR surgery OR surgeries OR surgical* OR visit* OR doctor* OR physician*) ) ) OR SU ( ( (emergency OR emergencies OR disaster*) ) NOT ( (emergency OR emergencies) N2 (accident OR care OR service* OR department* OR room OR rooms OR unit OR units OR ward OR wards OR hospital* OR medical* OR surgery OR surgeries OR surgical* OR visit* OR doctor* OR physician*) ) ) ) ) | Search modes - Boolean/Phrase | 684 |
| S2 | ( TI ( ((pandemic* OR "public health" OR "health authorit*" OR "health department*" OR "health education" OR "health promotion" OR "health protection" OR "health system*" OR "health unit*" OR community OR communities OR county OR counties OR municipal* OR neighborhood OR neighborhoods OR neighbourhood OR neighbourhoods OR ((region OR regions OR regional*) N3 health) OR parish OR parishes OR population OR populations OR (outbreak* N5 (diseas* OR COVID* OR SARS* OR "severe acute respiratory syndrome" OR ebola* OR H1N1* OR zika OR MERS OR "middle east respiratory syndrome")) OR epidemic*) and ((emergency OR emergencies OR disaster*) not ((emergency OR emergencies) N2 (accident OR care OR service* OR department* OR room OR rooms OR unit OR units OR ward OR wards OR hospital* OR medical* OR surgery OR surgeries OR surgical* OR visit* OR doctor* OR physician*)))) ) OR SU ( ((pandemic* OR "public health" OR "health authorit*" OR "health department*" OR "health education" OR "health promotion" OR "health protection" OR "health system*" OR "health unit*" OR community OR communities OR county OR counties OR municipal* OR neighborhood OR neighborhoods OR neighbourhood OR neighbourhoods OR ((region OR regions OR regional*) N3 health) OR parish OR parishes OR population OR populations OR (outbreak* N5 (diseas* OR COVID* OR SARS* OR "severe acute respiratory syndrome" OR ebola* OR H1N1* OR zika OR MERS OR "middle east respiratory syndrome")) OR epidemic*) and ((emergency OR emergencies OR disaster*) not ((emergency OR emergencies) N2 (accident OR care OR service* OR department* OR room OR rooms OR unit OR units OR ward OR wards OR hospital* OR medical* OR surgery OR surgeries OR surgical* OR visit* OR doctor* OR physician*)))) ) OR KW ( ((pandemic* OR "public health" OR "health authorit*" OR "health department*" OR "health education" OR "health promotion" OR "health protection" OR "health system*" OR "health unit*" OR community OR communities OR county OR counties OR municipal* OR neighborhood OR neighborhoods OR neighbourhood OR neighbourhoods OR ((region OR regions OR regional*) N3 health) OR parish OR parishes OR population OR populations OR (outbreak* N5 (diseas* OR COVID* OR SARS* OR "severe acute respiratory syndrome" OR ebola* OR H1N1* OR zika OR MERS OR "middle east respiratory syndrome")) OR epidemic*) and ((emergency OR emergencies OR disaster*) not ((emergency OR emergencies) N2 (accident OR care OR service* OR department* OR room OR rooms OR unit OR units OR ward OR wards OR hospital* OR medical* OR surgery OR surgeries OR surgical* OR visit* OR doctor* OR physician*)))) ) ) OR ( TI ( ("public health emergency" OR "public health emergencies" OR ((pandemic* OR "public health" OR epidemic* OR (diseas* N2 (outbreak* OR infect* OR emerging OR communicable)) OR COVID* OR SARS* OR "severe acute respiratory syndrome" OR ebola* OR H1N1* OR zika OR MERS OR "middle east respiratory syndrome" OR epidemic*) N5 (emergency OR emergencies OR disaster*))) ) OR SU ( ("public health emergency" OR "public health emergencies" OR ((pandemic* OR "public health" OR epidemic* OR (diseas* N2 (outbreak* OR infect* OR emerging OR communicable)) OR COVID* OR SARS* OR "severe acute respiratory syndrome" OR ebola* OR H1N1* OR zika OR MERS OR "middle east respiratory syndrome" OR epidemic*) N5 (emergency OR emergencies OR disaster*))) ) OR KW ( ("public health emergency" OR "public health emergencies" OR ((pandemic* OR "public health" OR epidemic* OR (diseas* N2 (outbreak* OR infect* OR emerging OR communicable)) OR COVID* OR SARS* OR "severe acute respiratory syndrome" OR ebola* OR H1N1* OR zika OR MERS OR "middle east respiratory syndrome" OR epidemic*) N5 (emergency OR emergencies OR disaster*))) ) ) | Search modes - Boolean/Phrase | 1,614 |
| S3 | S1 OR S2 | Search modes - Boolean/Phrase | 1,738 |
| S4 | ( DE "CIVIL defense" OR DE "EMERGENCY management" OR DE "DISASTER medicine" OR DE "HEALTH planning" OR DE "SOCIAL planning" OR DE "STRATEGIC planning" OR DE "CAPACITY building" OR DE "GOVERNMENT programs" OR DE "PILOT projects" OR DE "GOVERNMENT regulation" OR DE "MEDICAL policy" OR DE "GOVERNMENT policy" OR DE "FORECASTING" OR DE "RESOURCE allocation" OR DE "SECURITY systems" OR DE "PUBLIC safety" OR DE "EMERGENCY communication systems" OR DE "EMERGENCY sanitation" OR DE "CRISIS management" OR DE "DISASTER resilience" OR DE "EMERGENCY management" OR DE "SOCIAL adjustment" OR DE "RESILIENCE (Personality trait)" OR DE "ORGANIZATIONAL resilience" OR DE "RESILIENT design" OR DE "POSTTRAUMATIC growth" OR DE "INTERNATIONAL relief" OR DE "DISASTER relief" OR DE "DISASTER relief -- Government policy") ) OR ( TI ( (prepar* OR plan OR plans OR plann* OR manag* OR train* OR resilien* OR coping OR cope OR respon* OR ready OR readiness OR recover* OR rebuild* OR "build* back" OR "bounc* back" OR countermeasure* OR "public health preparedness" OR "public health emergency preparedness" OR "pandemic preparedness" OR "pandemic emergency preparedness") ) OR SU ( (prepar* OR plan OR plans OR plann* OR manag* OR train* OR resilien* OR coping OR cope OR respon* OR ready OR readiness OR recover* OR rebuild* OR "build* back" OR "bounc* back" OR countermeasure* OR "public health preparedness" OR "public health emergency preparedness" OR "pandemic preparedness" OR "pandemic emergency preparedness") ) OR KW ( (prepar* OR plan OR plans OR plann* OR manag* OR train* OR resilien* OR coping OR cope OR respon* OR ready OR readiness OR recover* OR rebuild* OR "build* back" OR "bounc* back" OR countermeasure* OR "public health preparedness" OR "public health emergency preparedness" OR "pandemic preparedness" OR "pandemic emergency preparedness") ) ) | Search modes - Boolean/Phrase | 99,624 |
| S5 | ( (DE "INFECTION control" OR DE "INFECTION prevention") ) OR ( TI ( ((((emergencies OR emergency) NOT ((emergency OR emergencies) N2 (accident OR care OR service* OR department* OR room OR rooms OR unit OR units OR ward OR wards OR hospital* OR medical* OR surgery OR surgeries OR surgical* OR visit* OR doctor* OR physician*))) OR pandemic* OR epidemic* OR disaster* OR "mass casualt*" OR COVID* OR SARS* OR "severe acute respiratORy syndrome" OR future OR post-emergenc* OR post-pandemic* OR post-covid* OR post-sars* OR (diseas* N2 (infect* OR emerging OR communicable))) N3 (prepar* OR plan OR plans OR plann* OR manag* OR train* OR resilien* OR coping OR cope OR respon* OR ready OR readiness OR recover* OR rebuild* OR "build* back" OR "bounc* back" OR countermeasure*)) ) OR KW ( ((((emergencies OR emergency) NOT ((emergency OR emergencies) N2 (accident OR care OR service* OR department* OR room OR rooms OR unit OR units OR ward OR wards OR hospital* OR medical* OR surgery OR surgeries OR surgical* OR visit* OR doctor* OR physician*))) OR pandemic* OR epidemic* OR disaster* OR "mass casualt*" OR COVID* OR SARS* OR "severe acute respiratORy syndrome" OR future OR post-emergenc* OR post-pandemic* OR post-covid* OR post-sars* OR (diseas* N2 (infect* OR emerging OR communicable))) N3 (prepar* OR plan OR plans OR plann* OR manag* OR train* OR resilien* OR coping OR cope OR respon* OR ready OR readiness OR recover* OR rebuild* OR "build* back" OR "bounc* back" OR countermeasure*)) ) OR SU ( ((((emergencies OR emergency) NOT ((emergency OR emergencies) N2 (accident OR care OR service* OR department* OR room OR rooms OR unit OR units OR ward OR wards OR hospital* OR medical* OR surgery OR surgeries OR surgical* OR visit* OR doctor* OR physician*))) OR pandemic* OR epidemic* OR disaster* OR "mass casualt*" OR COVID* OR SARS* OR "severe acute respiratORy syndrome" OR future OR post-emergenc* OR post-pandemic* OR post-covid* OR post-sars* OR (diseas* N2 (infect* OR emerging OR communicable))) N3 (prepar* OR plan OR plans OR plann* OR manag* OR train* OR resilien* OR coping OR cope OR respon* OR ready OR readiness OR recover* OR rebuild* OR "build* back" OR "bounc* back" OR countermeasure*)) ) ) | Search modes - Boolean/Phrase | 12,612 |
| S6 | S4 OR S5 | Search modes - Boolean/Phrase | 99,730 |
| S7 | (DE "EVALUATION" OR DE "EVALUATION research (Social action programs)" OR DE "RETROSPECTIVE studies" OR DE "BENCHMARKING (Management)" OR DE "OUTCOME assessment (Social services)" OR DE "PERFORMANCE evaluation" OR DE "SYSTEM analysis" OR DE "QUALITATIVE research" OR DE "EVALUATION research" OR DE "EVALUATION methodology" OR DE "PROGRAM effectiveness (Education)" OR DE "PROGRAM improvement (Education)" OR DE "EDUCATIONAL evaluation" OR DE "TOTAL quality management" OR DE "QUALITY control" OR DE "MANAGEMENT audit" OR DE "DELPHI method" OR DE "NEEDS assessment" OR DE "KEY performance indicators (Management)" OR DE "STANDARDS") | Search modes - Boolean/Phrase | 3,751 |
| S8 | ( TI ( ((evaluat* OR apprais* OR assess* OR benchmark* OR improv* OR enhanc* OR quality OR ineffective* OR efficac* OR effective* OR outcome* OR audit* OR analyz* OR analys* OR ((preparedness OR response) N3 (capacity OR capacities OR capability OR capabilities)) OR (quality N3 (assess* OR improv* OR manag* OR control* OR assur*))) N3 (reporting OR report OR measur* OR metric* OR score* OR scoring OR scorecard* OR standard* OR target OR targets OR "COVID-score*" OR "report card*" OR perform* OR quality OR ineffective* OR efficac* OR effective* OR outcome* OR lesson* OR framework* OR indicator* OR index OR indexes OR instrument OR instruments OR inventories OR inventory OR registries OR registry OR implement* OR initiativ* OR intervention OR interventions OR model OR models OR tool OR tools OR theor* OR schema* OR pilot* OR plan OR plans OR planning OR policy OR policies OR principles OR program OR programs OR programme OR programmes OR roadmap* OR "road map*" OR strategy OR strategies)) ) OR KW ( ((evaluat* OR apprais* OR assess* OR benchmark* OR improv* OR enhanc* OR quality OR ineffective* OR efficac* OR effective* OR outcome* OR audit* OR analyz* OR analys* OR ((preparedness OR response) N3 (capacity OR capacities OR capability OR capabilities)) OR (quality N3 (assess* OR improv* OR manag* OR control* OR assur*))) N3 (reporting OR report OR measur* OR metric* OR score* OR scoring OR scorecard* OR standard* OR target OR targets OR "COVID-score*" OR "report card*" OR perform* OR quality OR ineffective* OR efficac* OR effective* OR outcome* OR lesson* OR framework* OR indicator* OR index OR indexes OR instrument OR instruments OR inventories OR inventory OR registries OR registry OR implement* OR initiativ* OR intervention OR interventions OR model OR models OR tool OR tools OR theor* OR schema* OR pilot* OR plan OR plans OR planning OR policy OR policies OR principles OR program OR programs OR programme OR programmes OR roadmap* OR "road map*" OR strategy OR strategies)) ) OR SU ( ((evaluat* OR apprais* OR assess* OR benchmark* OR improv* OR enhanc* OR quality OR ineffective* OR efficac* OR effective* OR outcome* OR audit* OR analyz* OR analys* OR ((preparedness OR response) N3 (capacity OR capacities OR capability OR capabilities)) OR (quality N3 (assess* OR improv* OR manag* OR control* OR assur*))) N3 (reporting OR report OR measur* OR metric* OR score* OR scoring OR scorecard* OR standard* OR target OR targets OR "COVID-score*" OR "report card*" OR perform* OR quality OR ineffective* OR efficac* OR effective* OR outcome* OR lesson* OR framework* OR indicator* OR index OR indexes OR instrument OR instruments OR inventories OR inventory OR registries OR registry OR implement* OR initiativ* OR intervention OR interventions OR model OR models OR tool OR tools OR theor* OR schema* OR pilot* OR plan OR plans OR planning OR policy OR policies OR principles OR program OR programs OR programme OR programmes OR roadmap* OR "road map*" OR strategy OR strategies)) ) ) OR ( TI ( ((lesson* N2 learn*) OR "after-action review*" OR "after action review*" OR "in-action review*" OR "in action review*" OR scorecard* OR "COVID-score*" OR "report card*") ) OR SU ( ((lesson* N2 learn*) OR "after-action review*" OR "after action review*" OR "in-action review*" OR "in action review*" OR scorecard* OR "COVID-score*" OR "report card*") ) OR KW ( ((lesson* N2 learn*) OR "after-action review*" OR "after action review*" OR "in-action review*" OR "in action review*" OR scorecard* OR "COVID-score*" OR "report card*") ) ) | Search modes - Boolean/Phrase | 5,680 |
| S9 | S7 OR S8 | Search modes - Boolean/Phrase | 8,857 |
| S10 | S3 AND S6 AND S9 | Search modes - Boolean/Phrase | 72 |
| S11 | S3 AND S6 AND S9 | Limiters - Publication Date: 20170601-; Language: English Search modes - Boolean/Phrase | 21 |

**Table 5. Elsevier Scopus Search Query**

| # | Searches | Results |
| --- | --- | --- |
| 1 | TITLE-ABS-KEY ( ( ( pandemic* OR "public health" OR "health authorit*" OR "health department*" OR "health education" OR "health promotion" OR "health protection" OR "health system*" OR "health unit*" OR community OR communities OR county OR counties OR municipal* OR neighborhood OR neighborhoods OR neighbourhood OR neighbourhoods OR ( ( region OR regions OR regional* ) W/3 health ) OR parish OR parishes OR population OR populations OR ( outbreak* W/5 ( diseas* OR covid* OR sars* OR "severe acute respiratory syndrome" OR ebola* OR h1n1* OR zika OR mers OR "middle east respiratory syndrome" ) ) OR epidemic* ) AND ( ( emergency OR emergencies OR disaster* ) AND NOT ( ( emergency OR emergencies ) W/2 ( accident OR care OR service* OR department* OR room OR rooms OR unit OR units OR ward OR wards OR hospital* OR medical* OR surgery OR surgeries OR surgical* OR visit* OR doctor* OR physician* ) ) ) ) ) | 85,991 |
| 2 | TITLE-ABS-KEY ( "public health emergency" OR "public health emergencies" OR ( ( pandemic* OR "public health" OR epidemic* OR ( diseas* W/2 ( outbreak* OR infect* OR emerging OR communicable ) ) OR covid* OR sars* OR "severe acute respiratory syndrome" OR ebola* OR h1n1* OR zika OR mers OR "middle east respiratory syndrome" OR epidemic* ) W/5 ( emergency OR emergencies OR disaster* ) ) ) | 18,812 |
| 3 | #1 OR #2 | 92,447 |
| 4 | TITLE ( prepar* OR plan OR plans OR plann* OR manag* OR train* OR resilien* OR coping OR cope OR respon* OR ready OR readiness OR recover* OR rebuild* OR "build* back" OR "bounc* back" OR countermeasure* OR "public health preparedness" OR "public health emergency preparedness" OR "pandemic preparedness" OR "pandemic emergency preparedness" ) | 4,006,144 |
| 5 | TITLE-ABS-KEY ( ( ( ( emergencies OR emergency ) AND NOT ( ( emergency OR emergencies ) W/2 ( accident OR care OR service* OR department* OR room OR rooms OR unit OR units OR ward OR wards OR hospital* OR medical* OR surgery OR surgeries OR surgical* OR visit* OR doctor* OR physician* ) ) ) OR pandemic* OR epidemic* OR disaster* OR "mass casualt*" OR covid* OR sars* OR "severe acute respiratory syndrome" OR future OR post-emergenc* OR post-pandemic* OR post-covid* OR post-sars* OR ( diseas* W/2 ( infect* OR emerging OR communicable ) ) ) W/3 ( prepar* OR plan OR plans OR plann* OR manag* OR train* OR resilien* OR coping OR cope OR respon* OR ready OR readiness OR recover* OR rebuild* OR "build* back" OR "bounc* back" OR countermeasure* ) ) | 291,685 |
| 6 | #4 OR #5 | 4,190,538 |
| 7 | TITLE( ( evaluat* OR apprais* OR assess* OR benchmark* OR improv* OR enhanc* OR quality OR ineffective* OR efficac* OR effective* OR outcome* OR audit* OR analyz* OR analys* OR ( ( preparedness OR response ) W/3 ( capacity OR capacities OR capability OR capabilities ) ) OR ( quality W/3 ( assess* OR improv* OR manag* OR control* OR assur* ) ) ) W/3 ( reporting OR report OR measur* OR metric* OR score* OR scoring OR scorecard* OR standard* OR target OR targets OR "COVID-score*" OR "report card*" OR perform* OR quality OR ineffective* OR efficac* OR effective* OR outcome* OR lesson* OR framework* OR indicator* OR index OR indexes OR instrument OR instruments OR inventories OR inventory OR registries OR registry OR implement* OR initiativ* OR intervention OR interventions OR model OR models OR tool OR tools OR theor* OR schema* OR pilot* OR plan OR plans OR planning OR policy OR policies OR principles OR program OR programs OR programme OR programmes OR roadmap* OR "road map*" OR strategy OR strategies ) ) | 2,586,038 |
| 8 | TITLE-ABS ( ( lesson* W/2 learn* ) OR "after-action review*" OR "after action review*" OR "in-action review*" OR "in action review*" OR scorecard* OR "COVID-score*" OR "report card*" ) | 114,697 |
| 9 | #7 OR #8 | 2,692,207 |
| 10 | #3 AND #6 AND #9 | 3,364 |
| 11 | PUBYEAR > 2016 AND LANGUAGE ( english ) | 16,942,123 |
| 12 | #10 AND #11 | 1,711 |
| 13 | AFFILCOUNTRY ( "Austria" OR "Baltic State*" OR "Belgium" OR "Chile" OR "Colombia" OR "Costa Rica" OR "Czech Republic" OR "Developed Countr*" OR "Estonia" OR "Europe" OR "Australia" OR "Canada" OR "Denmark" OR "France" OR "Germany" OR "Italy" OR "Japan" OR "Korea" OR "Norway" OR "United Kingdom" OR "England" OR "United States" OR "Finland" OR "Greece" OR "Hungary" OR "Iceland" OR "Ireland" OR "Scotland" OR "Wales" OR "Israel" OR "Latvia" OR "Lithuania" OR "Luxembourg" OR "Mexico" OR "Netherlands" OR "New Zealand" OR "North America" OR "Poland" OR "Portugal" OR "Slovakia" OR "Slovenia" OR "Spain" OR "Sweden" OR "Switzerland" OR "Turkey" OR "Undefined" ) | 53,436,518 |
| 14 | #12 AND #13 | 1,247 |
| 15 | AFFILCOUNTRY ( "Afghanistan" OR "Argentina" OR "Western Asia" OR "Asia" OR "Bahrain" OR "Bangladesh" OR "Belize" OR "Bhutan" OR "Bolivia" OR "Brazil" OR "Central America" OR "Developing Countr*" OR "Ecuador" OR "El Salvador" OR "Africa" OR "Central Asia" OR "Northern Asia" OR "Southeastern Asia" OR "Caribbean" OR "China" OR "India" OR "Far East" OR "French Guiana" OR "Guatemala" OR "Guyana" OR "Honduras" OR "Iran" OR "Iraq" OR "Jordan" OR "Kuwait" OR "Latin America" OR "Lebanon" OR "Middle East" OR "Mongolia" OR "Nepal" OR "Nicaragua" OR "Oman" OR "Pakistan" OR "Panama" OR "Paraguay" OR "Peru" OR "Qatar" OR "Saudi Arabia" OR "South America" OR "Sri Lanka" OR "Suriname" OR "Syria" OR "Taiwan" OR "Turkey" OR "United Arab Emirates" OR "Uruguay" OR "Venezuela" OR "Yemen" ) | 16,767,790 |
| 16 | #12 AND NOT #15 | 1,294 |
| 17 | #14 OR #16 | 1,457 |
| 18 | #17 AND NOT INDEX (medline OR embase) | 730 |

**Grey literature search queries**

**Search concept legend:**

- Emergency/pandemic/covid-19/public health emergencies**:** pandemic OR coronavirus OR covid OR sars ; emergency OR pandemic OR epidemic OR coronavirus OR covid OR sars ; emergency OR pandemic OR epidemic ; coronavirus OR covid OR sar ; "public health" OR "infectious disease*” ; "public health" OR community OR "health system" emergency
- Post-emergency/post-pandemic/post-covid: after OR aftermath OR post emergency OR pandemic OR epidemic OR coronavirus OR covid OR sars ; post-emergency OR post-pandemic OR post-covid ; post-emergency OR post-pandemic OR post-covid
- Preparedness/management/response/resilience/recovery: "emergency preparedness" OR "emergency plan*" OR "emergency management" OR "emergency response" OR "system preparedness" OR "community preparedness" OR "health preparedness" ; preparedness OR management OR response ; planning OR preparedness OR response OR readiness OR management ; "emergency preparedness" OR "emergency plan" OR "emergency management" OR "emergency response" OR "system preparedness" ; "pandemic preparedness" OR "pandemic plan" OR "pandemic management" OR "pandemic response" ; planning OR preparedness OR response OR readiness OR management ; preparedness OR management OR response ; resilience OR recovery OR build back OR bounce back OR rebuild OR coping OR cope ; preparedness OR management OR response
- Evaluation/assessment: "lessons learned" OR "after-action review" OR "after action review" OR evaluation OR assess OR performance OR quality
- Indicator/measure: indicator OR measure OR metric OR scorecard Or reporting
- Frameworks/models: framework OR initiative OR model OR pilot OR roadmap OR "road map" OR strategy OR strategies

**Grey literature databases searched:**

Custom search engines (n=7)

- [Search for Canadian Federal and Provincial Emergency Management Agencies](https://cse.google.com/cse/publicurl?cx=011843882881040462305:nh8ooaox6pu)
- [Search Ontario's Public Health Units](https://cse.google.com/cse?cx=005247453031710928926:wszuea0acm8)
- [Search Canadian Health Departments and Agencies](https://cse.google.com/cse?cx=54dae8807550ea08a)
- [Search U.S. State Government Websites](https://cse.google.com/cse?cx=f7f9d691627b6ed48)
- [Search for International Public Health Resources](https://cse.google.com/cse?cx=b15f5d17bb6cc614d)
- [Search .Gov/.Org/.Edu domains (these are U.S. domains)](https://cse.google.com/cse/publicurl?cx=006912983573246070439:dbea-6lyv1k)
- [Google Canada](https://www.google.ca/)

Site specific searching: core public health agencies /government websites (n=20)

- BC Centre for Disease Control: site:.bccdc.ca/ [enter search query]
- Centers for Disease Control and Prevention: site:.cdc.gov/
- European Centre for Disease Prevention and Control: site:.ecdc.europa.eu/
- European Union: site:.europa.eu/
- Federal Emergency Management Agency site:.fema.gov/
- Government of Australia: site:.gov.au
- Government of Canada: site:.gc.ca/
- Government of Ireland: site:.gov.ie
- Government of New Zealand: site:.govt.nz
- Government of the United Kingdom: site:.gov.uk
- Government of the United States: site:.gov/
- Institut national de santé publique du Québec: site:.inspq.qc.ca/
- International Association of Emergency Managers site:.iaem.com/
- National Emergency Management Association [site:.nemaweb.org/](http://www.nemaweb.org/)
- Organisation for Economic Co-operation and Development: site:.oecd-ilibrary.org/
- Public Health Agency of Canada: site:.phac-aspc.gc.ca/
- United Nations: site:.un.org/
- World Association for Disaster and Emergency Medicine [site:.wadem.org/](https://wadem.org/)
- World Bank: site:.worldbank.org/
- World Health Organization: site:.who.int/

**Table 6. Grey literature search queries**

| Search # | Search Query |
| --- | --- |
| **1** | pandemic OR coronavirus OR covid OR sars "emergency preparedness" OR "emergency plan*" OR "emergency management" OR "emergency response" OR "system preparedness" OR "community preparedness" OR "health preparedness" evaluation OR assess OR performance OR quality indicator OR measure OR metric OR scorecard |
| **2** | emergency OR pandemic OR epidemic OR coronavirus OR covid OR sars preparedness OR management OR response evaluation OR assess OR performance OR quality framework OR initiative OR model OR pilot OR roadmap OR "road map" OR strategy OR strategies |
| **3** | emergency OR pandemic OR epidemic planning OR preparedness OR response OR readiness OR management evaluation OR assess OR performance OR quality indicator OR measure OR metric OR scorecard |
| **4** | coronavirus OR covid OR sars planning OR preparedness OR response OR readiness OR management evaluation OR assess OR performance OR quality indicator OR measure OR metric OR scorecard |
| **5** | "public health" OR "infectious disease*" "emergency preparedness" OR "emergency plan" OR "emergency management" OR "emergency response" OR "system preparedness" evaluation OR assess OR performance OR quality indicator OR measure OR metric OR scorecard OR reporting |
| **6** | "pandemic preparedness" OR "pandemic plan" OR "pandemic management" OR "pandemic response" evaluation OR assess OR performance OR quality indicator OR measure OR metric OR scorecard OR reporting |
| **7** | "public health" OR community OR "health system" emergency planning OR preparedness OR response OR readiness OR management evaluation OR assess OR performance OR quality indicator OR measure OR metric OR scorecard |
| **8** | post-emergency OR post-pandemic OR post-covid preparedness OR management OR response evaluation OR assess OR performance OR quality indicator OR measure OR metric OR scorecard |
| **9** | post-emergency OR post-pandemic OR post-covid preparedness OR management OR response "lessons learned" OR "after-action review" OR "after action review" |
| **10** | post-emergency OR post-pandemic OR post-covid resilience OR recovery OR build back OR bounce back OR rebuild OR coping OR cope evaluation OR assess OR performance OR quality indicator OR measure OR metric OR scorecard |
| **11** | after OR aftermath OR post emergency OR pandemic OR epidemic OR coronavirus OR covid OR sars preparedness OR management OR response evaluation OR assess OR performance OR quality indicator OR measure OR metric OR scorecard |
